# Non-pharmaceutical behavioural measures for droplet-borne biological hazards prevention: Health-EDRM for COVID-19 (SARS-CoV-2) Pandemic

**DOI:** 10.1101/2020.05.29.20116475

**Authors:** Emily Ying Yang Chan, Tayyab Salim Shahzada, Tiffany Sze Tung Sham, Caroline Dubois, Zhe Huang, Sida Liu, Janice Ying-en Ho, Kevin KC Hung, Kwok Kin On, Ryoma Kayano, Rajib Shaw

**Author notes:** **Corresponding Author:** Professor Emily Ying Yang Chan Rm 308, 3/F, JC School of Public Health, Prince of Wales Hospital, Shatin, Hong Kong SAR.

## Abstract

**Introduction:** Non-pharmaceutical interventions to facilitate response to the COVID-19 pandemic, a disease caused by novel coronavirus SARS-CoV-2, are urgently needed. Using the WHO health emergency and disaster risk management (health-EDRM) framework, behavioural measures for droplet-borne communicable disease, with their enabling and limiting factors at various implementation levels were evaluated.

**Sources of data:** Keyword search was conducted in PubMed, Google Scholar, Embase, Medline, Science Direct, WHO and CDC online publication database. Using OCEBM as review criteria, 105 English-language articles, with ten bottom-up, non-pharmaceutical prevention measures, published between January 2000 and May 2020 were identified and examined.

**Areas of Agreement:** Evidence-guided behavioural measures against COVID-19 transmission for global at-risk communities are identified.

**Area of Concern:** Strong evidence-based systematic behavioural studies for COVID-19 prevention are lacking.

**Growing points:** Very limited research publications are available for non-pharmaceutical interventions to facilitate pandemic response.

**Areas timely for research:** Research with strong implementation feasibility that targets resource-poor settings with low baseline Health-EDRM capacity is urgently need.

## Introduction

Uncertainties in disease epidemiology, treatment and management in biological hazards have often urged policy makers and community health protection to revisit prevention approaches to maximise infection control and protection. The COVID-19 pandemic, a disease caused by novel coronavirus SARS-CoV-2, has pushed global governments and communities to revisit the appropriate non-pharmaceutical, health prevention measures in response to this unexpected virus outbreak^1^. The World Health Organisation (WHO) health emergency and disaster risk management (health-EDRM) framework refers to the structured analysis and management of health risks brought upon by emergencies and disasters. The framework focuses on prevention and risk mitigation through hazard and vulnerability reduction, preparedness, response and recovery measures^2^, and further calls attention to the significance of community involvement to counteract the potential negative impacts of hazardous events such as infectious disease outbreaks^2^. COVID-19 is defined as a biological hazard under the health-EDRM disaster classification^3^. While there is evidence for potential COVID-19 droplet transmission^4^, the WHO has suggested that airborne transmission may only be possible in certain circumstances^4^. Further evidence is needed to categorise it as an airborne disease specifically, per the framework.

Health-EDRM prevention measures can be classified into primary, secondary or tertiary levels^5^. Primary prevention mitigates the occurrence of illness through an emphasis on health promotion and education aimed at behavioural modification^6^; secondary prevention involves screening and infection identification; and tertiary prevention focuses on treatment. In the context of COVID-19, both secondary and tertiary preventive measures are complicated due to the high incidence of asymptomatic patients^7^; the lack of consensus and availability of specific treatment or vaccine^8^; and the added stress on the health system during a pandemic. Primary prevention that focuses on protecting an individual from contracting an infection^9^ is thus the most practical option. A comprehensive disaster management cycle (prevention, mitigation, preparedness, response and recovery) encompasses both top-down and bottom-up measures^10,11^. Top-down measures require well-driven bottom-up initiatives to successfully achieve primary prevention and effectively modify community behaviours^12^.

Based on the health-EDRM framework, which emphases the impact of context on efficacy of measure practices^3^, this paper examines available published evidence on primary prevention measures that might be adopted at the personal, household and community level for droplet-borne transmitted diseases, and enabling and limiting factors for each measure. Additionally, this paper reviews the strength of available scientific evidence for each of the behavioural changes measure which may reduce health risks.

## Methodology

A literature search was conducted in May 2020. English-language based, international peer-reviewed articles, online reports, electronic books and press releases published between January 2000 and May 2020 were identified. The snowballing search methodology was also utilised. Research databases examined in this study included *PubMed, Google Scholar, Embase, Medline and Science Direct*. WHO, United States Centers for Disease Control and Prevention (CDC) and other local-government publications and information outlets were also included. The following keywords or phrases were searched for: ‘health-EDRM’. ‘COVID-19’ ‘risk-management’, ‘droplet transmission’, ‘global health’, ‘epidemiology’, ‘virus’, ‘prevention’, ‘coronavirus treatment’, ‘severe acute respiratory syndrome’, ‘disinfection’, ‘transmission’ ‘SARS’ ‘SARSCoV-2’, ‘coronaviruses’, ‘enveloped viruses’, ‘virus stability’, ‘COVID-19 stability’, ‘virus transmission’, ‘host responses to virus’, ‘2019-nCOV’. ‘SARS-CoV-2 entry points’, ‘respiratory virus’, ‘air pollution, ‘handwashing’, ‘pollution mask’, ‘face masks’, ‘infection risk reduction’, ‘rural handwashing’, ‘face touching’, ‘coughing and sneezing’, ‘virus outbreak’, ‘mass gatherings’, ‘biocidal agents virus’, ‘utensil sharing risk’, ‘cutlery sharing risk’, ‘respiratory hygiene’, ‘toilet plume’, ‘phone hygiene’. ‘COVID-19 policies’, ‘sodium hypochlorite disinfection’, ‘alcohol disinfection’, ‘carbon footprint’, ‘water supply scarcity’, ‘influenza’, ‘swine flu’, ‘social isolation’, ‘quarantine’, ‘social distancing’, ‘open defecation’, ‘WHO COVID-19’, ‘CDC COVID-19’, ‘COVID-19 advice’, ‘respiratory emission’, ‘ethics COVID-19’, ‘hygiene education’. Primary prevention measures as well as risk factors for infectious disease transmission were reviewed in order to generate ten core preventative measures for discussion. The avoidance of cutlery sharing, for example, was generated after determining it as a highly preventable risk for infectious disease transmission. Three independent reviewers engaged in the review and agreed on the final research used.

The literature was categorised according to the Oxford Centre for Evidence-Based Medicine (OCEBM) Levels of Evidence (Fig. 1)^13^, which systemises strength of evidence into levels, based on process of study design and methodology.

**Fig 1.**
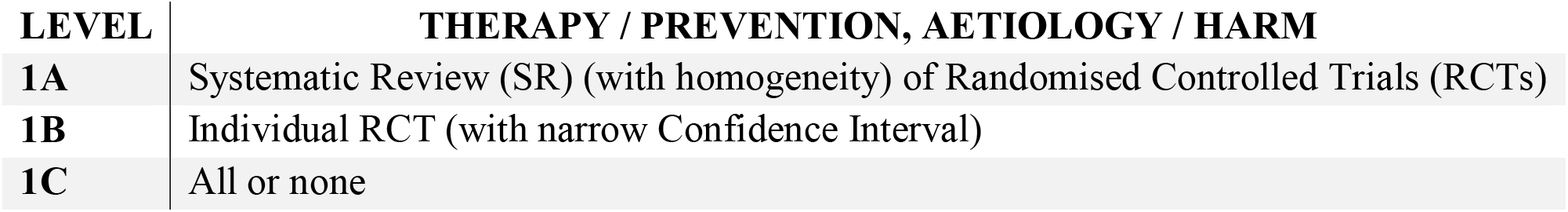
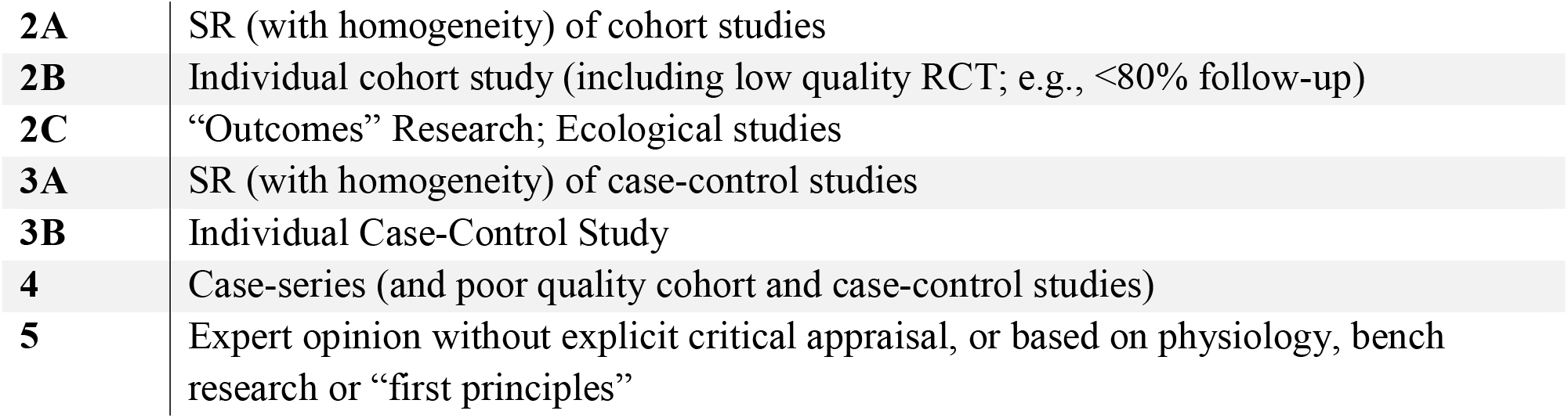
The Oxford Centre for Evidence-Based Medicine (OCEBM) Levels of Evidence^13^

## Results

The search identified 105 relevant publications, all of which were reviewed and included in the results analysis. The search identified ten bottom-up, non-pharmaceutical, primary prevention measures, based on the health-EDRM framework. The review of evidence is disaggregated into the ten prevention measures.

Six *personal* protective practices (engage in regular handwashing, wear face mask, avoid touching the face, cover mouth and nose when coughing and sneezing, bring personal utensils for when dining out, closing toilet cover when flushing), two *household* practices (disinfect household surfaces, avoid sharing cutlery) and two *community* practices (avoid crowds and mass gatherings, avoid travel) were identified. *Tables 1a, 1b and 1c* highlight the potential health risk; desired behavioural changes; potential health co-benefits; enabling and limiting factors; and strength of evidence available in published literature with regards to these measures.

**Table 1a (Part 1):** Personal practices as preventive measure - risk; behavioural change; health co-benefits; enabling and limiting factors; and strength of evidence.

|  | Engage in Regular Handwashing | Wear Face Mask | Avoid Touching the Face |
| --- | --- | --- | --- |
| **Risk** | • COVID-19 is transmittable through respiratory fluid droplets<sup>4,14</sup>.<br>• Droplets can persist on hands and other surfaces<sup>15</sup>.<br>• Droplets may be transferred if hands are not disinfected. | • Respiratory droplets from other individuals and hand-to-face contacts can result in droplet intake through the nose and mouth<sup>16,17,4</sup>.<br>• Viruses have the potential to survive in the respiratory tract<sup>18</sup>. The virus may also enter through ocular means, although studies focusing specifically on COVID-19 are limited<sup>19</sup>.<br>• COVID-19 has an incubation period of as long as 19 days<sup>20</sup>; asymptomatic or mildly symptomatic individuals may spread the virus through coughing or sneezing.<br>• In 2010, WHO stated that where there is improper mask usage, risk may increase<sup>21</sup>. | • Recent research has suggested that nasal carriage<sup>22</sup> and ocular entry<sup>19</sup> are key alternative routes to oral entry into the respiratory tract for COVID-19.<br>• It has been demonstrated that COVID-19 can be detected on surfaces of plastic, stainless steel, copper and cardboard for up to 72 hours<sup>15</sup> after contamination. Hand-to-face contact following contact of public surfaces may pose a risk. |
| **Behavioural Change** | • Wash hands with soap<sup>23,24,25,26,27,28,20,29</sup> for a minimum of 20 seconds using a step-by-step guideline such as the WHO healthcare-based 11-step guideline<sup>30</sup>.<br>• Wash hands before eating, after bathroom usage, after mask removal et cetera.<br>• Practice alternative handwashing routines as long as they maintain the core principle of ensuring that the entire surface area of the hands is scrubbed<sup>31</sup>.<br>• Ensure commonly missed areas are washed, such as the thumbs and fingertips<sup>32,33,34,35</sup>. | • Wear surgical face masks<sup>36,37,38,39,28,20,29</sup> to create a physical barrier preventing the spread or intake of the virus-containing respiratory droplet (which are released by coughing or sneezing) through facial openings<sup>40</sup>.<br>• Wear face masks to minimise the touching of the nose and mouth as these can serve as transmission routes for COVID-19<sup>16,41,17,4</sup>.<br>• Use face masks correctly to ensure the best overall effectiveness, including one-time usage; limiting usage to one day; and avoid touching the surface to minimise risk of self-contamination<sup>42,43</sup>. | • Avoid touching the face to minimise the risk of COVID-19 contact through the body's main entry points for transmittable conditions<sup>44,36,27,28,29</sup>: the mouth, the eyes and the nose.<br>• Exercise increased awareness of this unwanted practice to minimise the risk of infection, as self-touching of the face may be spontaneous<sup>45,46</sup>. |
| **Co-benefit(s)** | • Prevention of other contact-transmissible diseases such as influenza<sup>47,48</sup>, to some extent, diarrhoea<sup>49,50</sup> and eye infections<sup>51</sup>.<br>• Potential for reduced infection transmission in community and household<sup>52</sup>. | • Protection against other microbes transmitted by respiratory droplets through the nose, mouth or eyes<sup>19,22</sup>.<br>• Protection from air pollutants and other air particles<sup>53,54</sup>, which could cause other respiratory conditions<sup>55</sup> such as asthma and lung cancer<sup>56,57</sup>. | • Minimises contracting diseases with similar transmission pathways such as influenza<sup>58,44</sup>.<br>• Reduce risk of transferring bacterial pathogens found on hands<sup>59</sup>. |
| **Enabling Factor(s)** | • Availability and affordability of running and sufficient water, soap and alcohol-based rubs. | • Access to effective face masks.<br>• Information about the correct use of face masks,<br>including proper disposal.<br>• Information about when to wear facemasks.<br>• Socio-cultural acceptance and habit on wearing face masks (global East vs. West) | • Effectiveness may be limited for infants, children, and others who do not have sufficient conscious control of body movement. |
| **Limiting Factor(s) and/or Alternative(s)** | • Alcohol-based formulas as alternative; efficacy in killing enveloped viruses has been demonstrated<sup>60</sup>.<br>• Use of ash and mud as an alternative in areas where there is no access to soap or alcohol-based rubs, Although these carry potential antimicrobial properties<sup>61</sup>, their efficacy in counteracting viral infections is not well-evidenced<sup>62</sup>.<br>• In areas with scarce running water, sharing and reusing of water or water-container elevates risk of transmission through droplets<sup>63</sup>. | • For those who cannot access surgical face masks, due to affordability, availability or otherwise, homemade masks<sup>64</sup> accompanied with the same hygienic measures can be considered<sup>65</sup>. | • Where face-touching is necessary or difficult to control, for example in infants or children, handwashing will be a more effective prevention measure. |
| **Strength of Evidence** | • Published evidence showed handwashing is a core community prevention measure for COVID-19 transmission.<br>• Handwashing communities display lower risks of developing transmittable diseases when compared to their non-handwashing counterparts, in both rural<sup>61</sup> and urban populations<sup>66</sup>.<br>• The measure is recommended by multiple governing bodies, including the WHO<sup>36</sup> and CDC<sup>37</sup>.<br>• Studies from Severe Acute Respiratory Syndrome (SARS), although not conclusive, are suggestive of handwashing as an effective measure<sup>67</sup>.<br>• 20-seconds may be considered a minimum duration given that time reductions, for example to 5 seconds<sup>68</sup> have been demonstrated as less effective.<br>• Soap and alcohol-based rubs are well-evidenced in their capability to interact with and degrade enveloped viruses<sup>38,60</sup>.<br>• Evidence of ash or mud-based alternatives as antimicrobials is limited; no concrete evidence<br>with respect to efficacy against COVID-19 or other viral infections. | • Multiple extensive studies on the similar SARS coronavirus concluded that there is evidence of effective transmission risk reduction<sup>69,23,24,25</sup>. One study states uncertainty over the effectiveness<sup>70</sup>.<br>• Success of Hong Kong and Taiwan's high compliance to mask-wearing practices<sup>71</sup> has been potentially reflected in the low rate of locally infected cases of COVID-19<sup>72</sup>, with both communities having reported periods of no new infections despite initial surges<sup>73</sup>.<br>• Used in conjunction with other practices such as social distancing, a model simulation demonstrate community-wide benefits of mask-wearing<sup>74</sup>. | • Such measures have been recommended for influenza in the past<sup>26</sup>.<br>• There is strong evidence of viral infections entering through the facial entry points and has been demonstrated for COVID-19<sup>22,19</sup>, although evidence for impact of face-touching in disease-transmission was not found.<br>• The stability of COVID-19 virus on certain surfaces has been evidenced. Similarly, previous studies have demonstrated the stability of other coronaviruses such as SARS, on such surfaces<sup>75</sup>. |

**Table 1a (Part 2):**
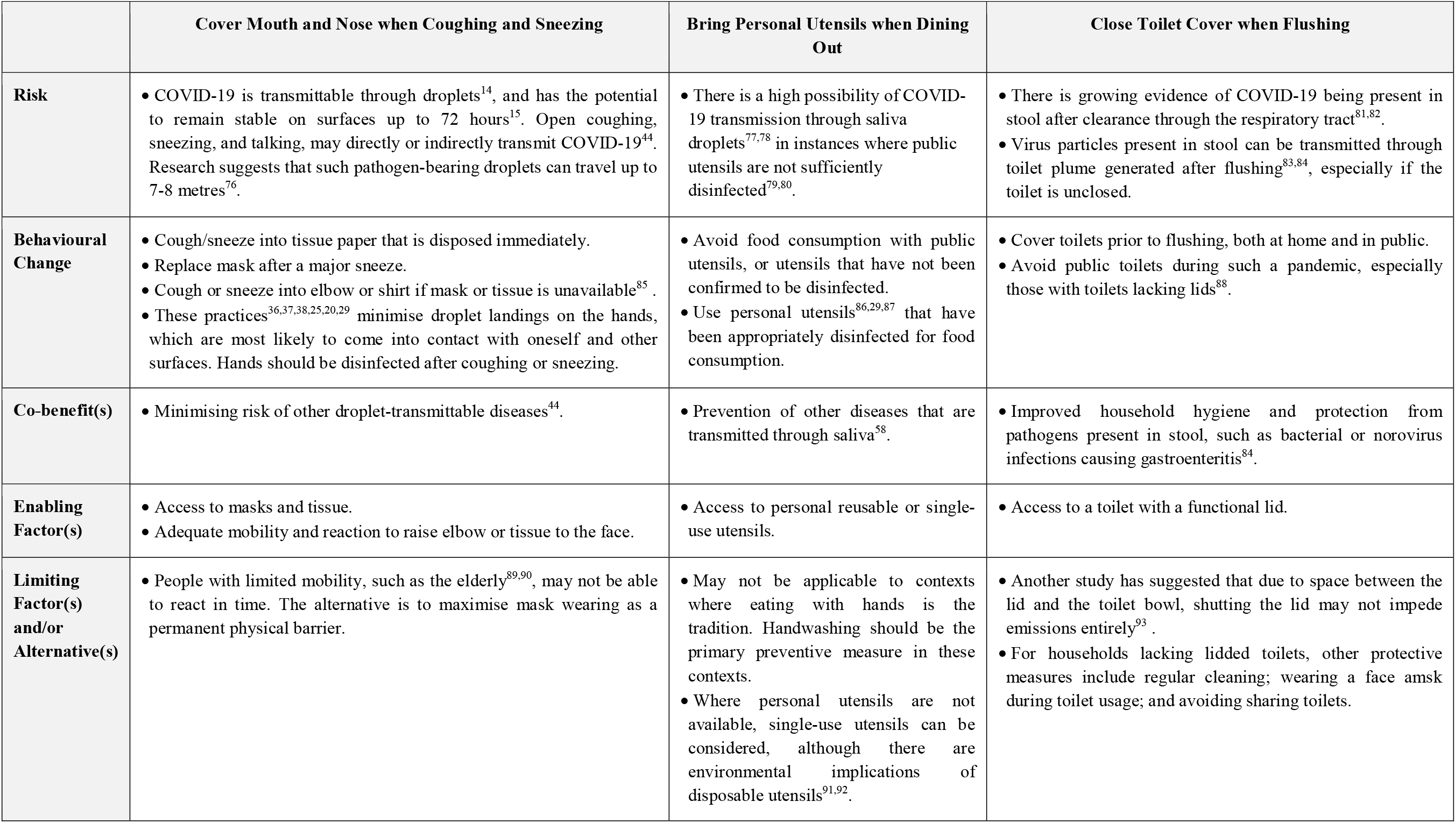
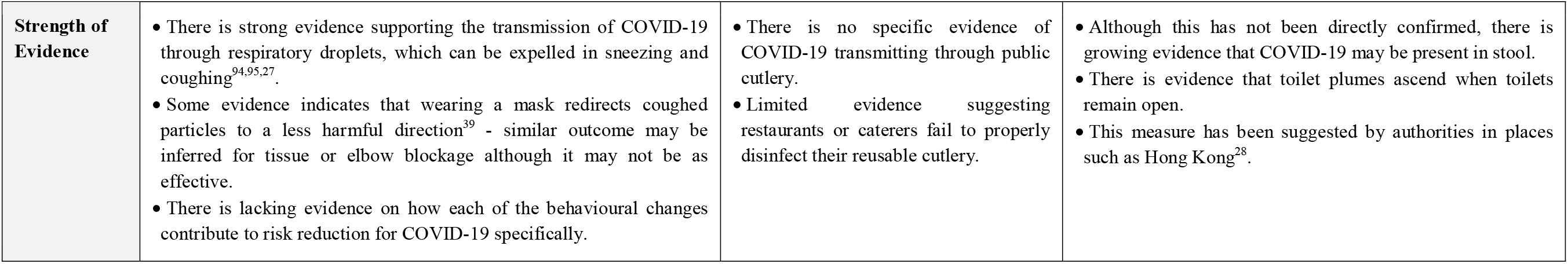
Personal practice as preventive measure - risk; behavioural change; health co-benefits; enabling and limiting factors; and strength of evidence.

**Table 1b:**
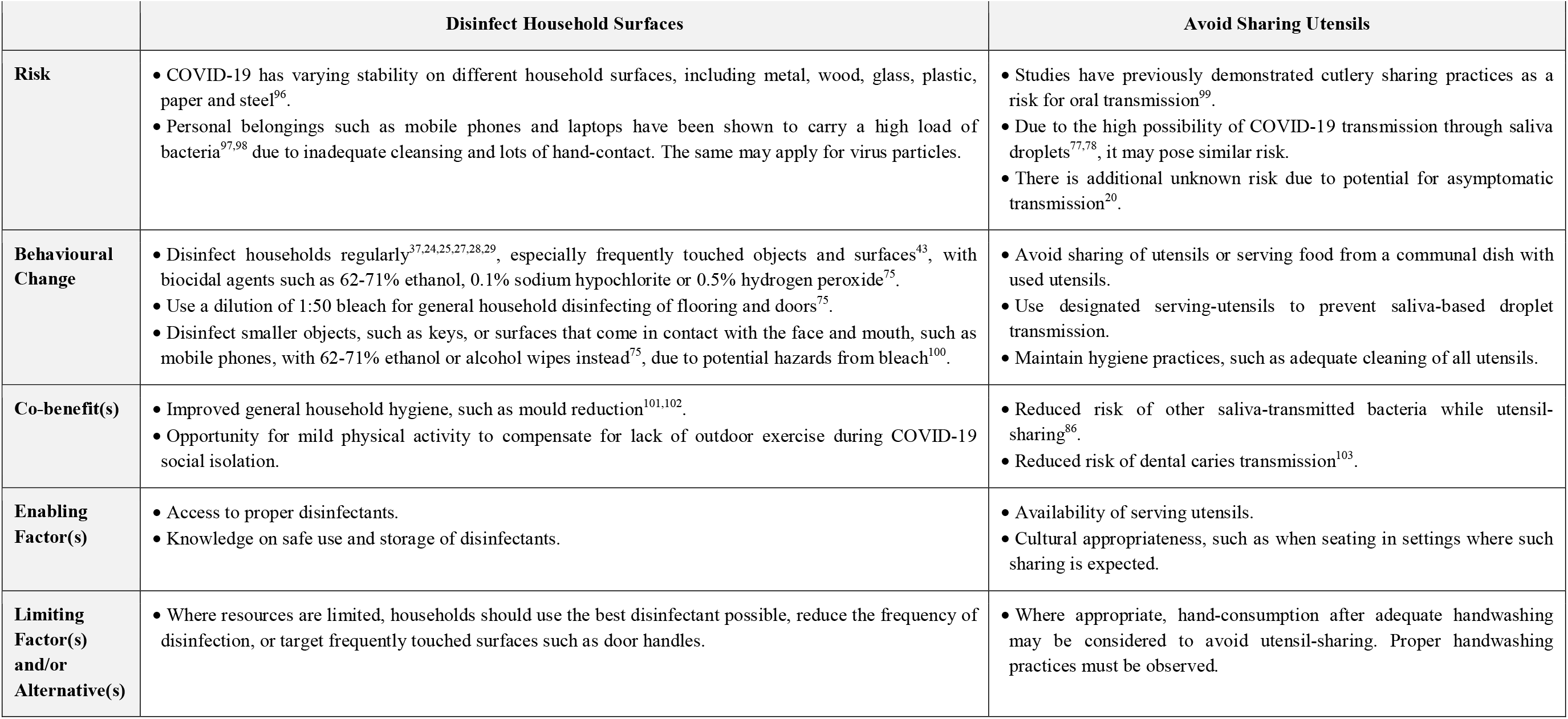
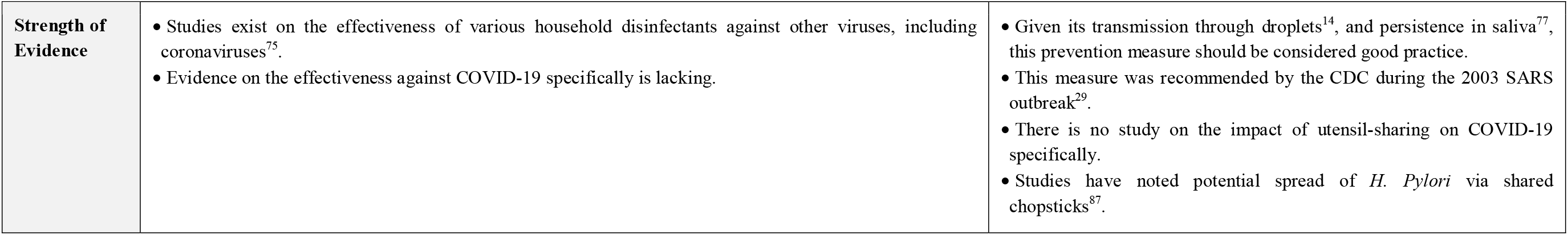
Household practices as preventive measure - risk; behavioural change; health co-benefits; enabling and limiting factors; and strength of evidence.

**Table 1b:**
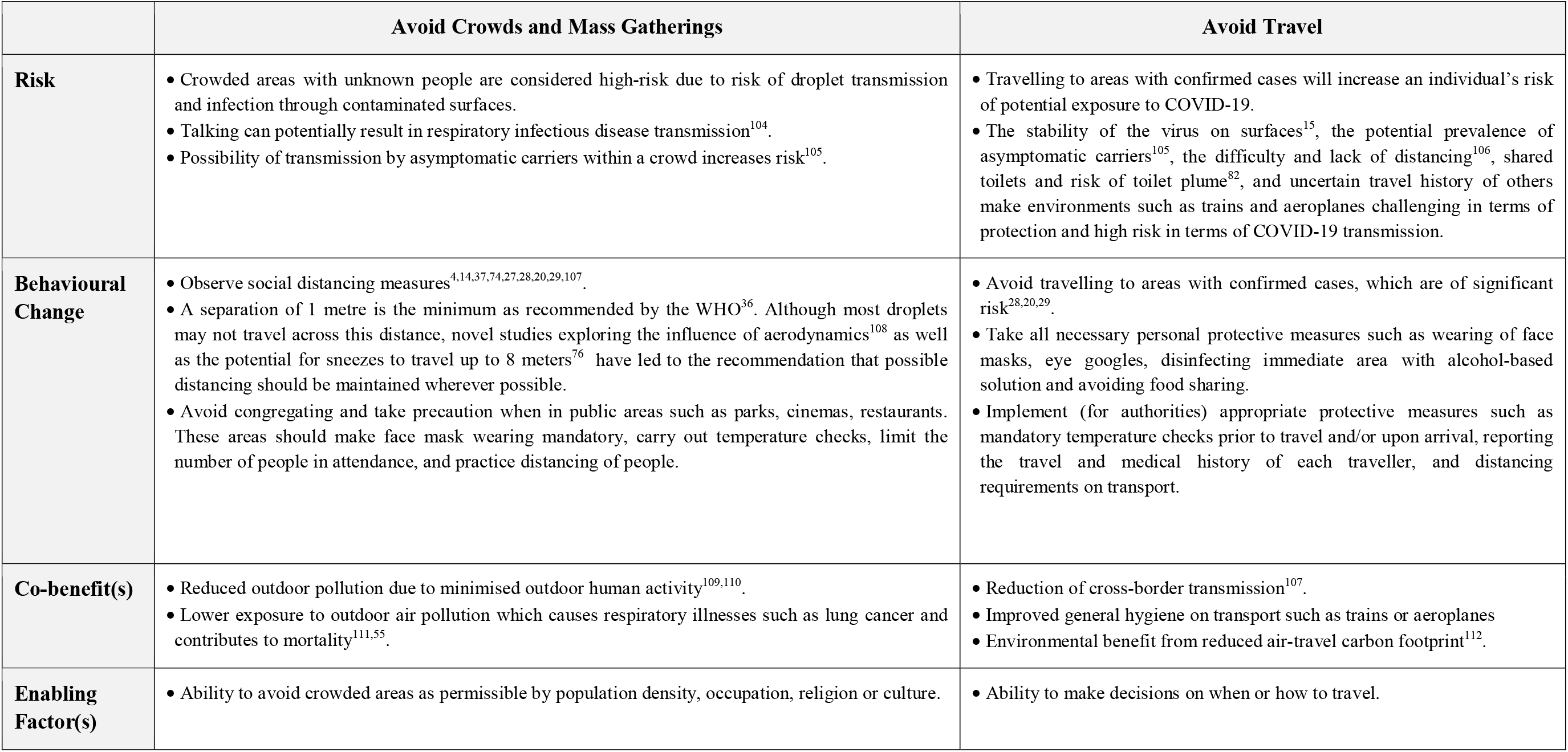
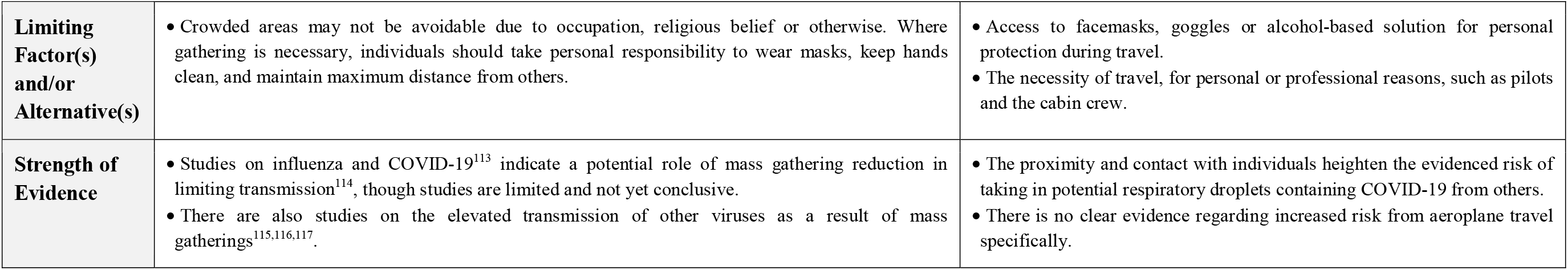
Community practice as preventive measure - risk; behavioural change; health co-benefits; enabling and limiting factors; and strength of evidence.

*Table 2* categorises all 105 reviewed publications according to the OCEBM Levels of Evidence^13^. Details each utilised reference can be found in *Appendix 1*. Quantity of evidence varies between each of ten measures reviewed, with handwashing and face mask practices having the most published resources available. Only a few systematic and longitudinal studies could be identified. Available evidence consists of predominantly level IV and V studies, and mainly of cross-sectional studies, guidelines and expert opinion.

**Table 2:** Number of referenced articles per measure by OCEBM categorisation level

| Category | Primary preventive measure | Number of referenced articles per OCEBM categorisation level |  |  |  |  |  |  |  |  |  |  |  |
| --- | --- | --- | --- | --- | --- | --- | --- | --- | --- | --- | --- | --- | --- |
|  |  | 1a | 1b | 1c | 2a | 2b | 2c | 3a | 3b | 4 | 5 | Others | Total |
| **Personal Practices** | Engage in Regular Handwashing | 4 | 5 | 0 | 0 | 1 | 0 | 2 | 2 | 3 | 17 | 0 | 34 |
|  | Wear Face Mask | 0 | 3 | 0 | 0 | 1 | 0 | 0 | 2 | 7 | 18 | 1 | 32 |
|  | Avoid Touching the Face | 0 | 0 | 0 | 0 | 2 | 0 | 0 | 0 | 1 | 10 | 0 | 13 |
|  | Cover Mouth and Nose when Coughing and Sneezing | 0 | 0 | 0 | 0 | 0 | 0 | 0 | 0 | 2 | 15 | 0 | 17 |
|  | Bring Personal Utensils for When Dining Out | 0 | 0 | 0 | 0 | 0 | 0 | 0 | 0 | 2 | 8 | 0 | 10 |
|  | Close Toilet Cover When Flushing | 0 | 0 | 0 | 0 | 0 | 0 | 0 | 0 | 2 | 6 |  | 8 |
| **Household Practices** | Disinfect Household Surfaces | 0 | 0 | 0 | 0 | 0 | 0 | 0 | 1 | 1 | 11 | 0 | 13 |
|  | Avoid Sharing Utensils | 0 | 0 | 0 | 0 | 1 | 0 | 0 | 0 | 4 | 4 | 0 | 9 |
| **Community Practices** | Avoid Crowds and Mass Gatherings | 0 | 0 | 0 | 0 | 0 | 0 | 0 | 0 | 2 | 21 | 0 | 23 |
|  | Avoid Travel | 0 | 0 | 0 | 0 |  | 0 | 0 | 0 | 3 | 6 | 0 | 9 |
| **Total** |  | 4 | 8 | 0 | 0 | 5 | 0 | 2 | 5 | 27 | 116 | 1 | 168* |

\* Some of the 105 publications are referenced against more than one of the ten primary preventive measures.

## Discussion

Evidence relating to ten health-EDRM behavioural measures for primary prevention against droplet-borne biological hazards were identified and reviewed. At the time of writing, there is an outstanding question as to whether COVID-19 is transmitted through droplet or aerosol in the community. The information referenced here is based on best available evidence, and will need to be updated as new studies and guidelines are published, and the understanding of the scientific community is enhanced. Although direct evidence on the efficacy of prevention measures against COVID-19 specifically is lacking due to the novelty of the disease, five behavioural measures were identified: regular handwashing; wearing of face masks; avoidance of face touching; covering during sneezing or coughing; and household disinfecting were identified. Five other potential behavioural measures were also identified through logical deductions from potential behavioural risks associated with transmission of diseases similar to COVID-19^75^. Utensil-related practices, in particular, were heavily limited in evidence to support their efficacy against viral infections.

The efficacy and success of the ten bottom-up primary prevention measures reviewed here are subject to specific enabling and limiting determinants, ranging from demographic (e.g. age, gender, education), socio-cultural, economic (e.g. financial accessibility to commodities), and knowledge (e.g. understanding of risk, equipment use). The viability and efficacy of each measure may be limited by determinants and constraints in different contexts. Resource-deprived areas may face constraints and reduced effectiveness of implementation, especially for measures that require preventive commodities such as face masks and household disinfectants. As such, special attention should be given to rural settings, informal settlements, and resource-deficit contexts where access to information and resources such as clean water supply are often limited^119,120^, and sanitation facilities are lacking^121^. For hygiene measures, different alternatives should be promoted and their relative scientific merits should be evaluated, such as the use of ash as an alternative to soap for handwashing^62^, or the efficacy of handwashing with alcohol sanitiser, which has been demonstrated in previously-published studies for H1N1^122^ and noroviruses^123^ but not yet for COVID-19. Meanwhile, for measures that have no direct alternatives available, it is important for authorities and policymakers to understand the capacity limitations of certain target groups, and provide additional support or put in place other preventive measures. In cases where material resources are scarce, the measures of awareness on sneezing and coughing etiquette as well as avoiding hand-to-face contact are the most convenient to adopt as they require little to no commodities. However, it should be well-noted that these measures are likely the most challenging in compliance and enforceability, as they rely on the modification of frequent and natural human behaviours whose modifications would require awareness and practice^45,46^. Furthermore, these can be challenging to implement in target groups with less capacity for health literacy and translation of education into practice, such as infants and elderly suffering from dementia. Cultural patterns can be associated with behavioural intentions, in the case of avoiding utensil-sharing during meals, enforcing change may be conflicted with cultural and traditional norms in Asia and certain European communities^124^.

Of the enabling factors documented for each proposed measure, shared enablers can be identified: accessibility and affordability of resources; related knowledge, awareness and understanding of risk; and associated top-down policy facilitation. Majority of personal and household practices heavily rely on access to resources, such as adequate water and soap supply for regular handwashing, quality face masks and household-disinfectants. Various theories of the ‘Knowledge, Attitudes, Practices’ model have assumed that individual knowledge enhancement will lead to positive behavioural changes^118^. Health measures targeting mask-wearing might aim to enhance (1) the individual’s risk perception, knowledge and awareness on protection effectiveness of masks, and how to properly wear a mask so that the prevention is most effective; (2) an individual or community’s attitude towards the practice of mask-wearing and encouraging compliance in the West, as studies demonstrate a relatively greater social stigmatisation towards mask-wearing amongst Westerners than East Asians^125^, and (3) normalising the practice of habitual mask-wearing. Such a conceptual framework should be utilized in the implementation of the health initiatives. In terms of overarching knowledge, health education on symptom-identification is also important, as seen on government platforms such as the CDC^37^. Enhancing health-seeking behaviour of potential carriers is critical to promoting a rapid response for quarantine or hospitalisation.

At the individual level, behavioural changes have different sustainability potentials and limitations. Measures can also result in unintended consequences, such as the improper disposal of face masks^126^ and the incorrect use of household disinfectants^127^ should be carefully monitored to maximise impact while minimising further health and safety risks. Top-down policy facilitation and strengthening of infrastructure will be essential for effective implementation. Top-down efforts in resource provision, such as the distribution of quality masks to all citizens by the government or similar authority^128^, enhance personal and household capacities to mitigate infection risks. On compliance, the effectiveness of community practices such as crowd and travel avoidance are highly dependent on the needs and circumstances of an individual and a community. More assertive top-down policies such as travel bans and social distancing rules may drive bottom-up initiatives within communities under legal deterrence^129^. However, in order to ensure population-level compliance to recommendations that have wide-ranging socioeconomic impact and involve more than a day-to-day behavioural change, careful risk and information communication is required, which takes in to consideration practical, legal and ethical aspects.

The strength of evidence available for each practice is dependent on multiple factors. In the case of a novel or emerging disease such as COVID-19, available evidence can be related specifically to the disease and pandemic, but some findings are deduced from studies on other similar viral infections and transmittable conditions, such as SARS or Influenza. Many interventions proposed by health authorities are not based on rigorous population-based longitudinal studies. While handwashing is well-regarded as a core measure by global and national public health agencies such as the WHO^36^ and CDC^37^ the chemical properties of eliminating enveloped viruses is understood^60,38^, specific studies on the practice’s efficacy and impact on COVID-19 transmission are lacking. Due to the uncertainties of disease pathology and epidemiology, effectiveness of behavioural measures against COVID-19 are far from conclusive. Other uncertainties are also reported on virus surface stability^15^ and if the efficacy of disinfectants against surface-stable viruses might vary with COVID-19^75^. Similar deductive evidence approaches from studies on other viruses have been utilised to judge the efficacy of face masks or the shutting of toilet lids^84,83^. Although published evidence suggested individual measures such as covering coughs and sneezes to be helpful against droplet transmissions^14^, further research is needed to understand the true efficacy of coverings such as masks, tissues or elbows as an adequate preventive measure against COVID-19. Given the rapid knowledge advancement and research updates related to COVID-19, further study updates will be warranted to identify the most appropriate behavioural measures to support bottom up biological hazard responses. Cost-effectiveness of the measures, their impact sustainability, co-benefits and risk implications on other sectors should also be examined and evaluated. Standardised studies across different contexts should be enhanced, for example conducting tests on the efficacy of different disinfectants or soaps under a standardised protocol. Such studies would increase evidence on individual and comparative efficacy.

The limitations in this review include language (English language only); database inclusion (grey literature not included); online accessibility of the article; and missed keywords. Publications documenting the experiences of traditional, non-English-speaking, rural communities during the COVID-19 pandemic might not be included in this review. Further research should review the measures’ efficacy in different contexts and make comparisons with their alternative measures. Specifically, alternative preventive measures that can be practiced in resource-poor, developing communities, whose health systems and economies generally suffer the greatest impact during pandemics are urgently needed. Increased understanding of how to effectively mitigate against biological hazards such as COVID-19 in various contexts will help communities prepare for future outbreaks and build disaster resilience in line with the recommendations from the health-EDRM framework.

Despite the constraints, this review has nevertheless identified common, relevant behavioural measures supported by best available evidence for the design and implementation of health policies that prevent droplet-borne biological hazards. Many of the measures recommended by authorities during the pandemic are based on best practice available rather than best available evidence. The possibility of conducting large cohort or randomised controlled studies is often complicated, and rather infeasible during a pandemic, as noted for face masks^130,131^. Further studies are needed to understand the efficacy of frequently proposed measures for transmission risk reduction. Nonetheless, each of the measures identified has scientific basis in mitigating the risk of droplet transmission^14^, either through personal measures such as handwashing, or community-based measures which aim to reduce person-to-person contact. It is important to explore the efficacy of alternatives, notably for transmission prevention and risk communication in low-resources or developing contexts where the capacity of the health system to mitigate and manage emergency events outbreaks are weak. For example, while face masks are understudied, the scientific study of cloth masks as an alternative is severely limited^65^, although recommended by the CDC^132^. Such alternative studies should expand to consider different cultures and contexts where different varieties of disinfectants, face masks and utensils may be used. There is also potential for comparative effectiveness studies to explore measures that provide the greatest transmission risk reduction at the lowest transaction cost to the individual and community and should thus be prioritised in low-resource contexts^133^.

## Conclusion

During the outbreak of a novel transmissible disease such as COVID-19, primary prevention is the strongest and most effective line of defence to reduce health risks when there is an absence of effective treatment or vaccine. COVID-19 is and will be subjected to ongoing research and scrutiny by global scientists, health professionals and policy makers. While research gaps remain on the efficacy of various health-EDRM prevention measures in risk reduction and transmission control of COVID-19, suboptimal scientific evidence does not negate the potential benefits arising from good hygiene practices, especially where the likelihood for negative outcome is minimal. Despite the lack of rigorous scientific evidence, the best available practice-based health education content, effective means of information dissemination, equitable access to resources, and monitoring of unintended consequences of the promoted measures, such as environmental pollution due to poor waste management, will be essential. A top-down approach should be multi-sectorial, bringing in policy makers with clinical, public health, environmental, and community management expertise to develop a coordinated and comprehensive approach in a globalised world.

## Data Availability

Data sharing is not applicable to this article as no new data were created or analyzed in this study.

## Appendix 1 Relevant Measure(s), Study Design, Relevant Key Finding(s) and/or Conclusion of Each Utilised Reference

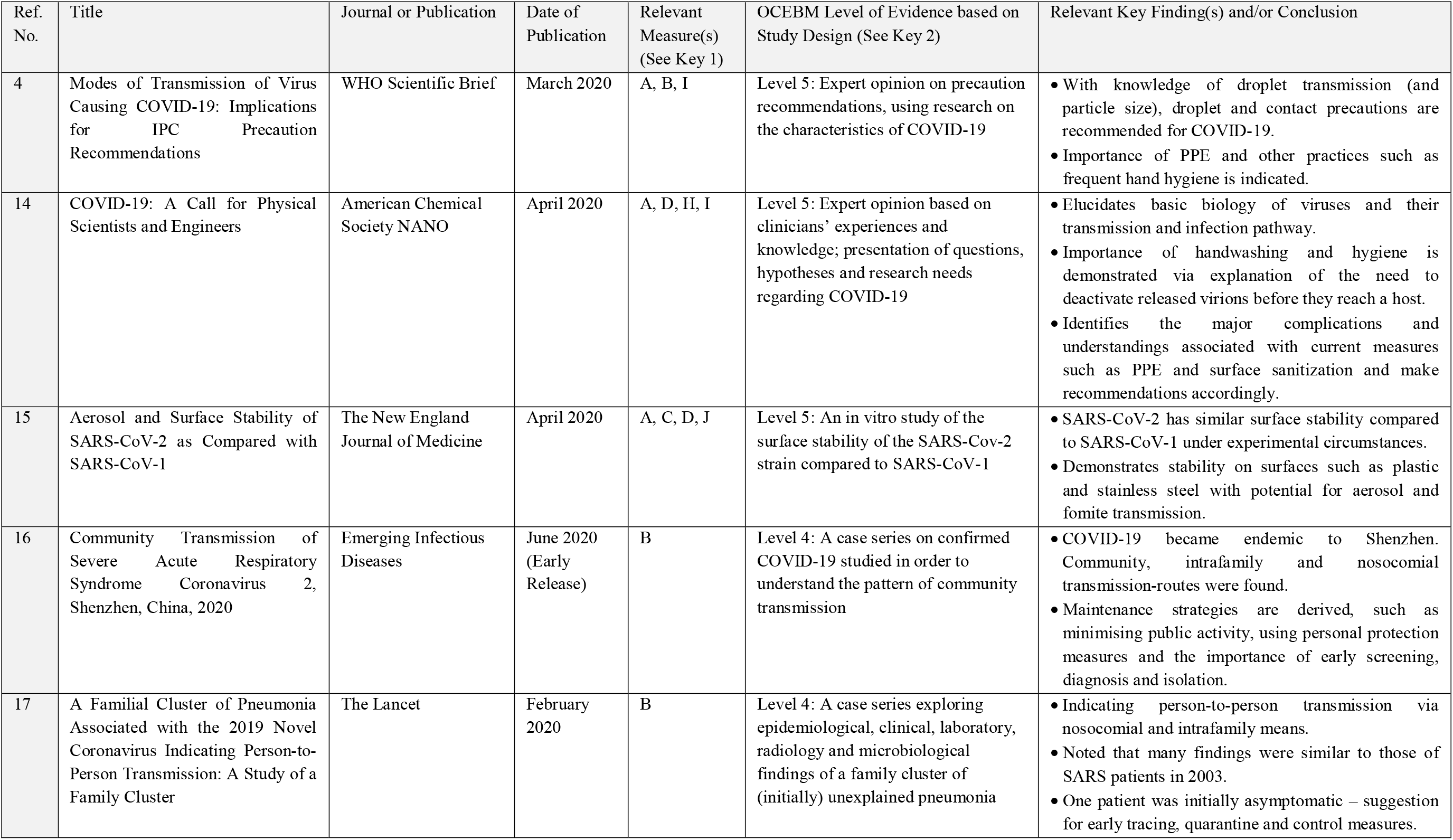
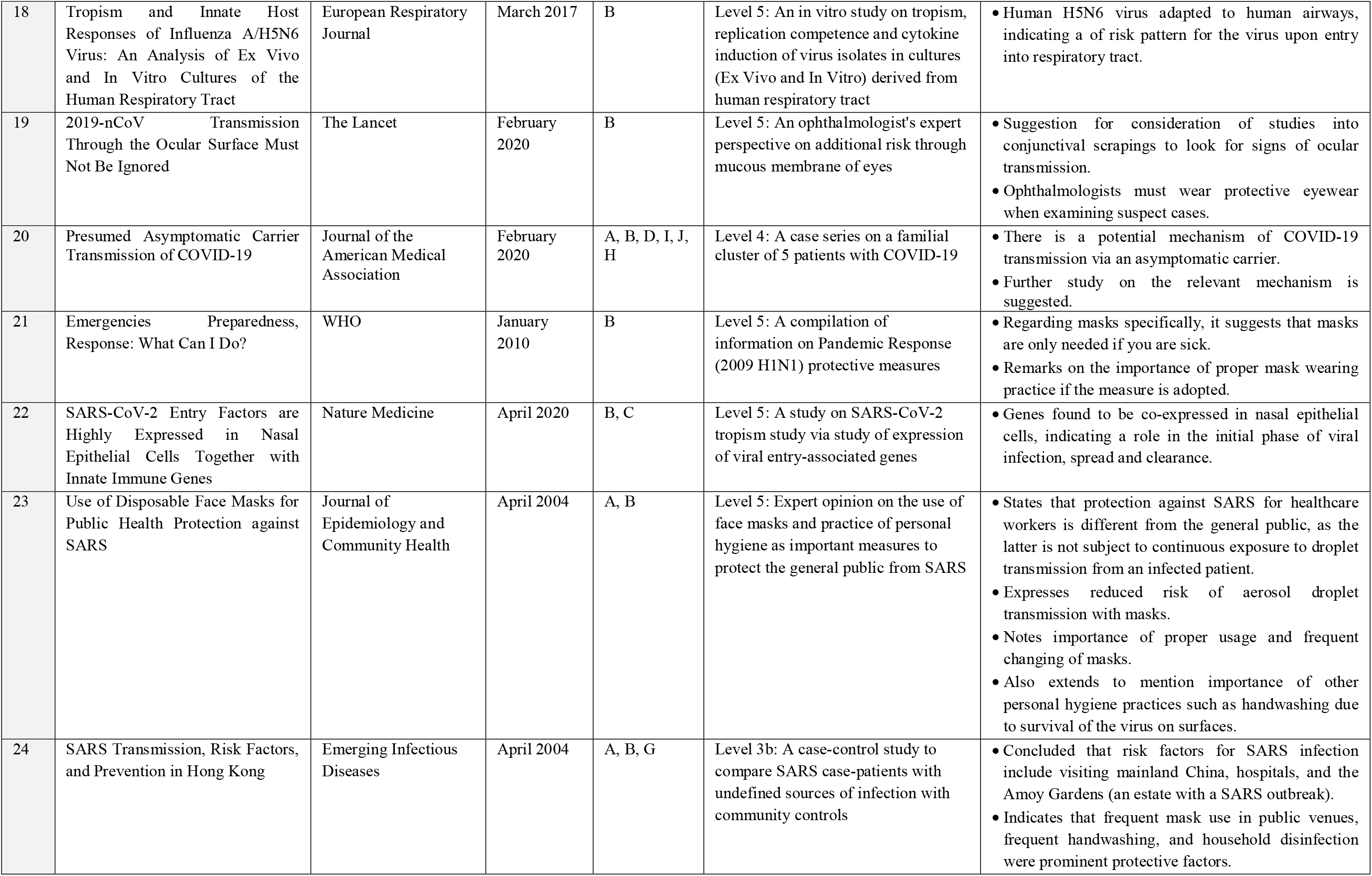
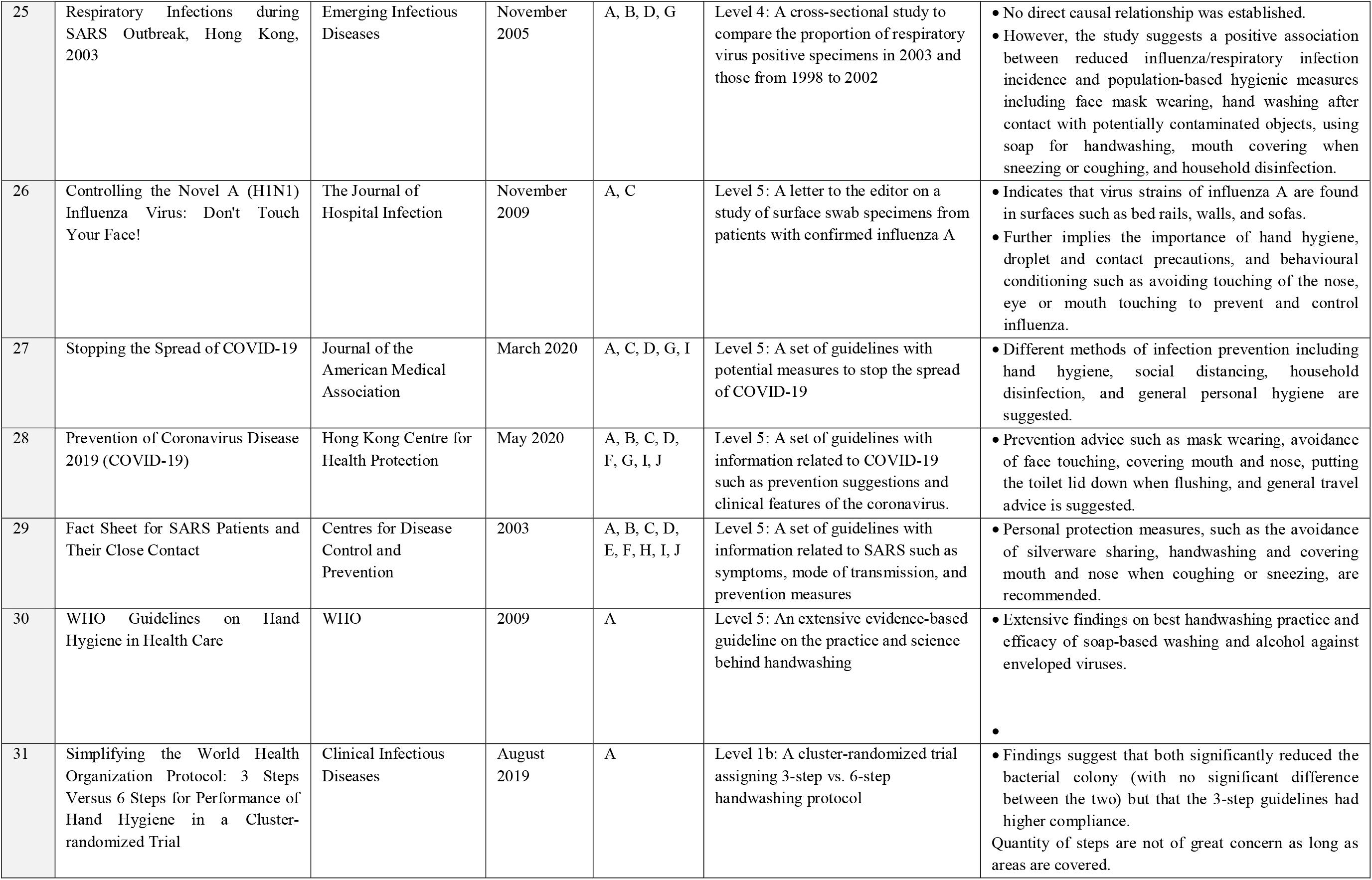
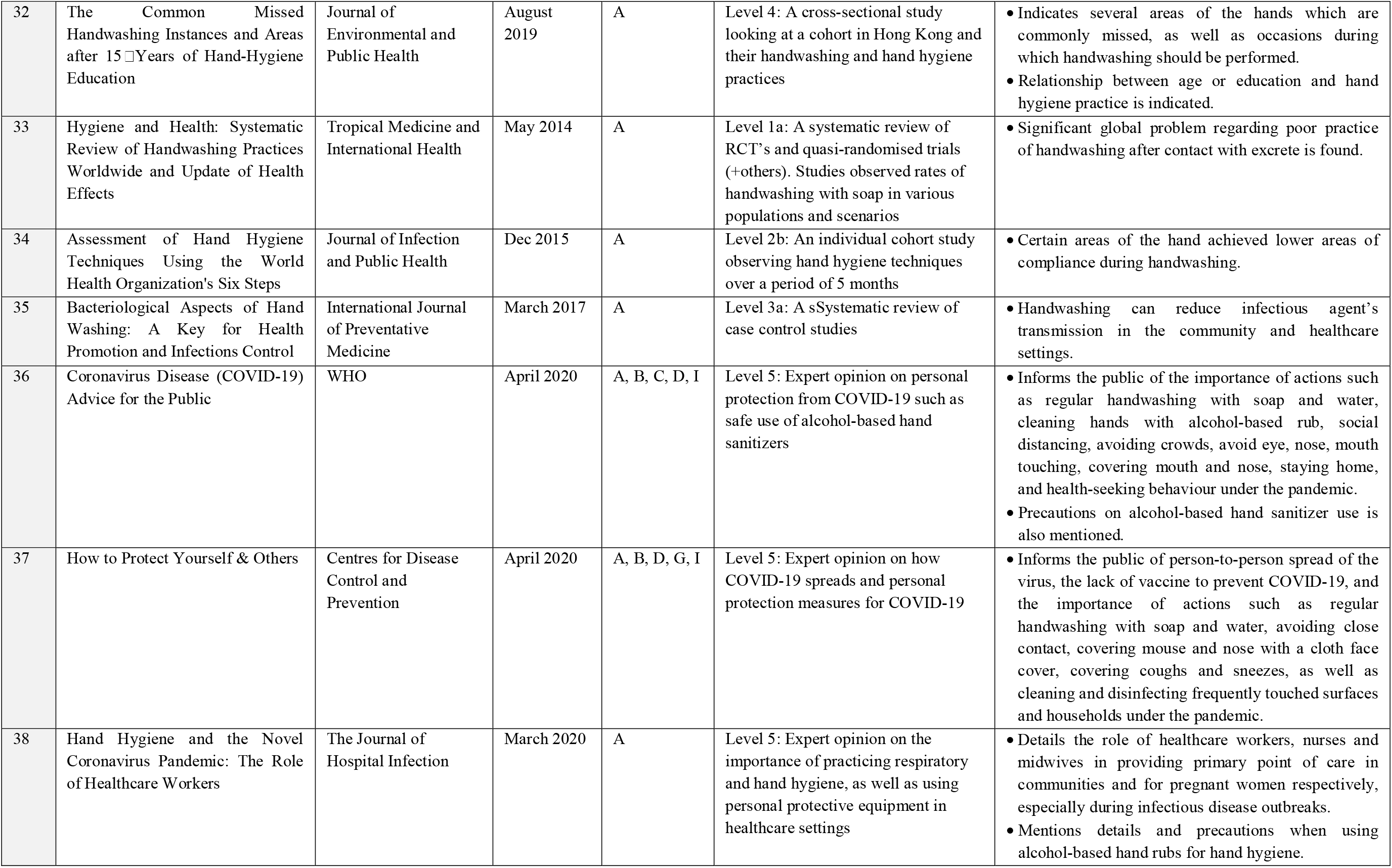
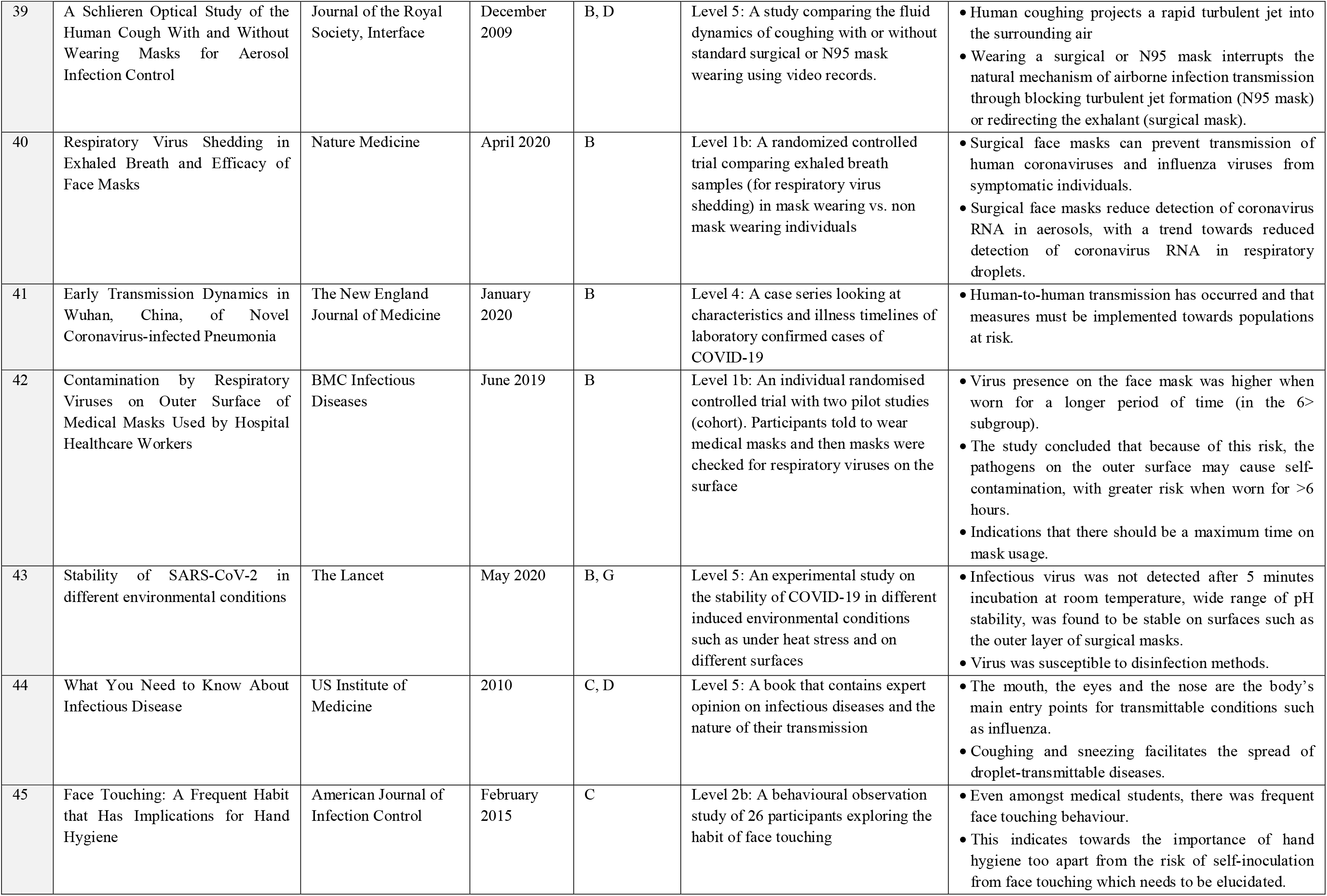
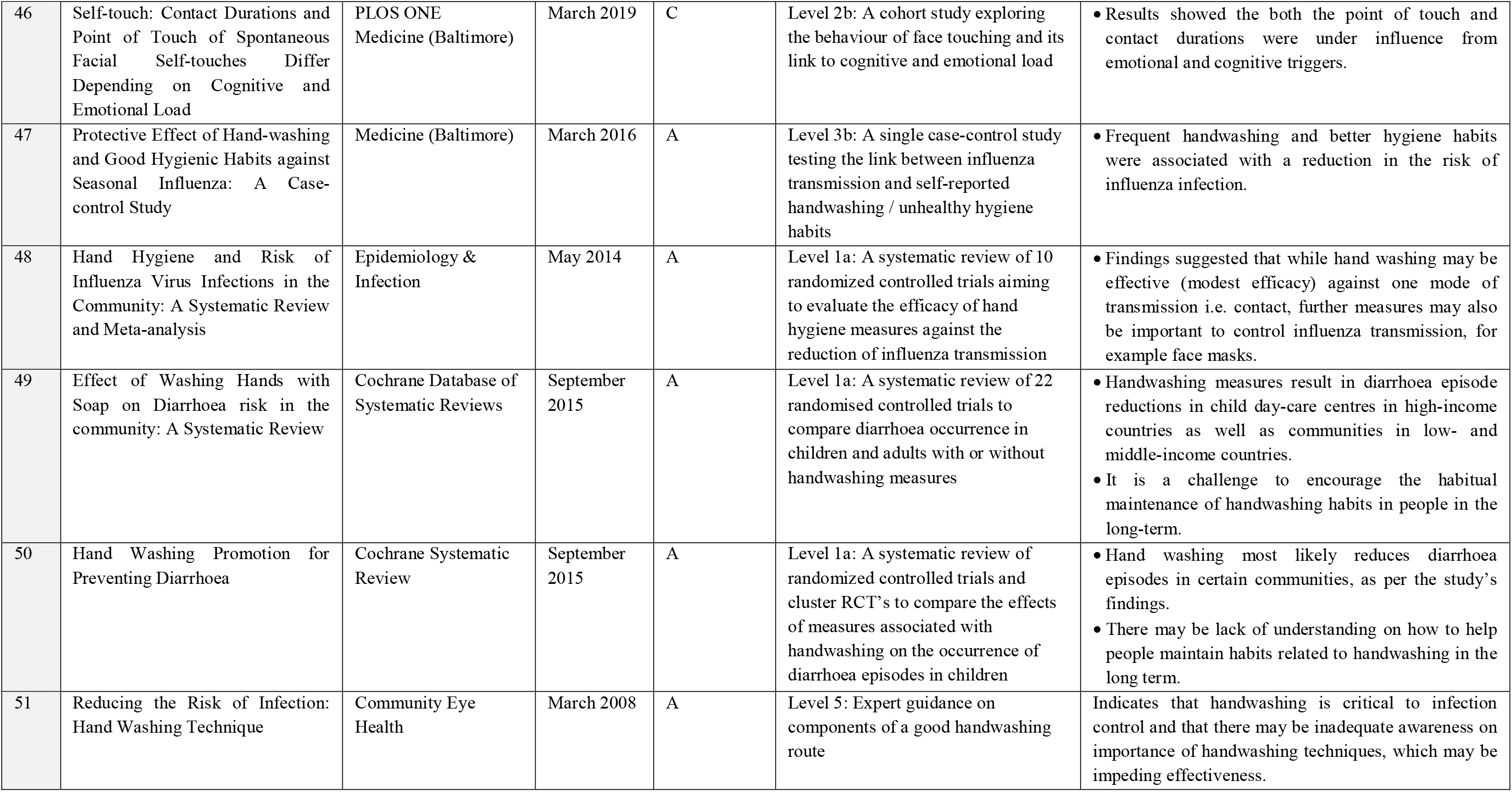
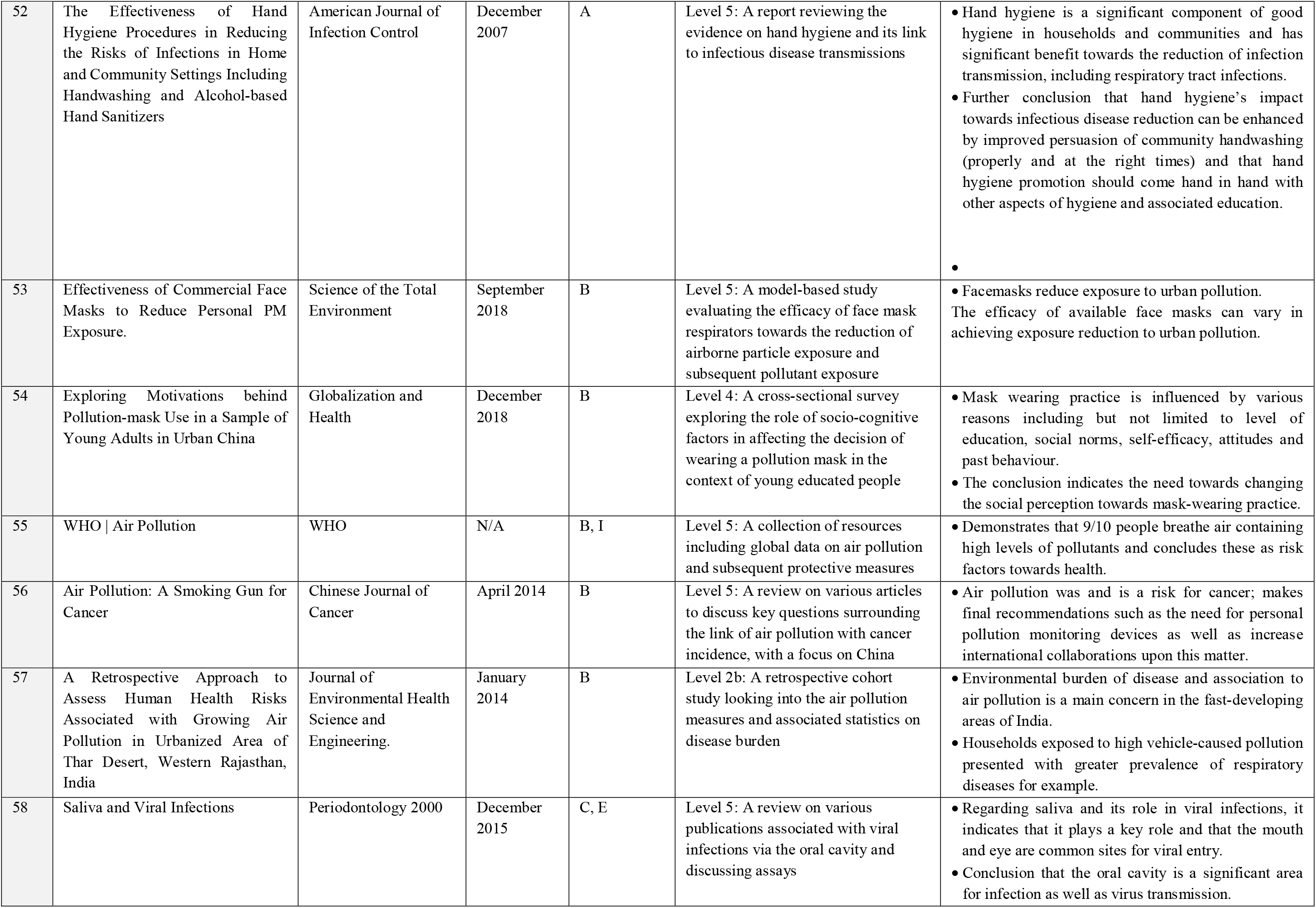
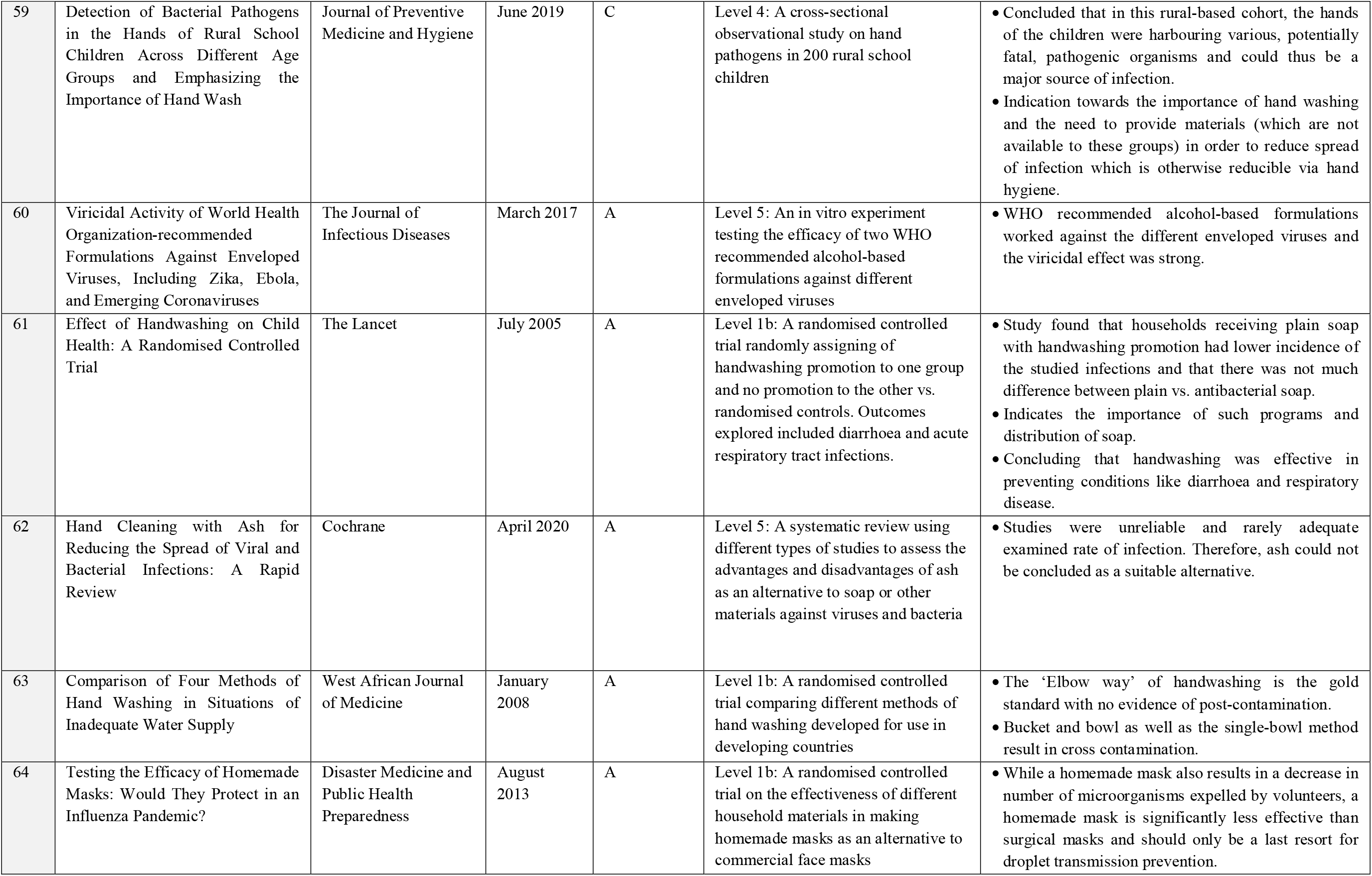
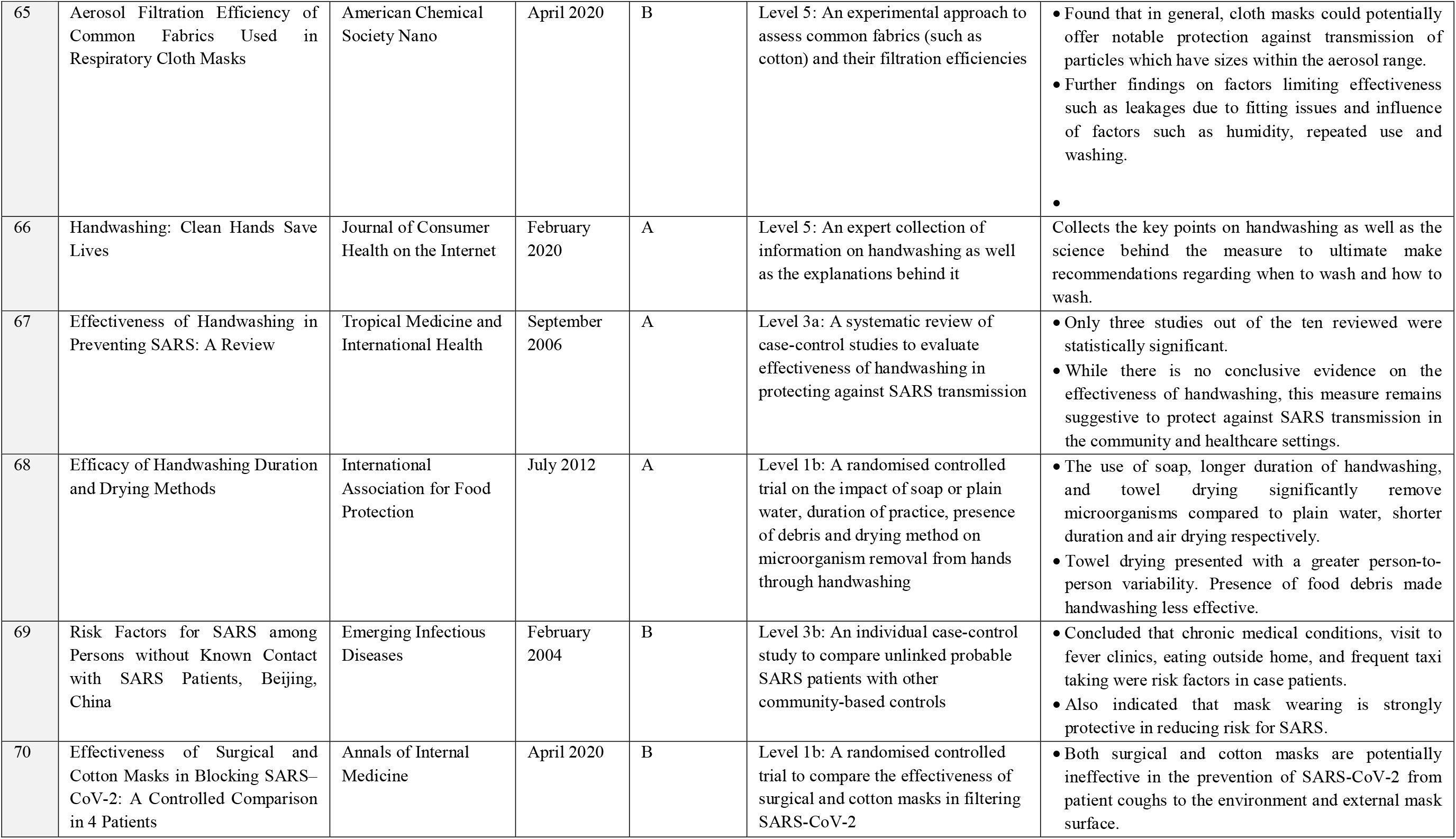
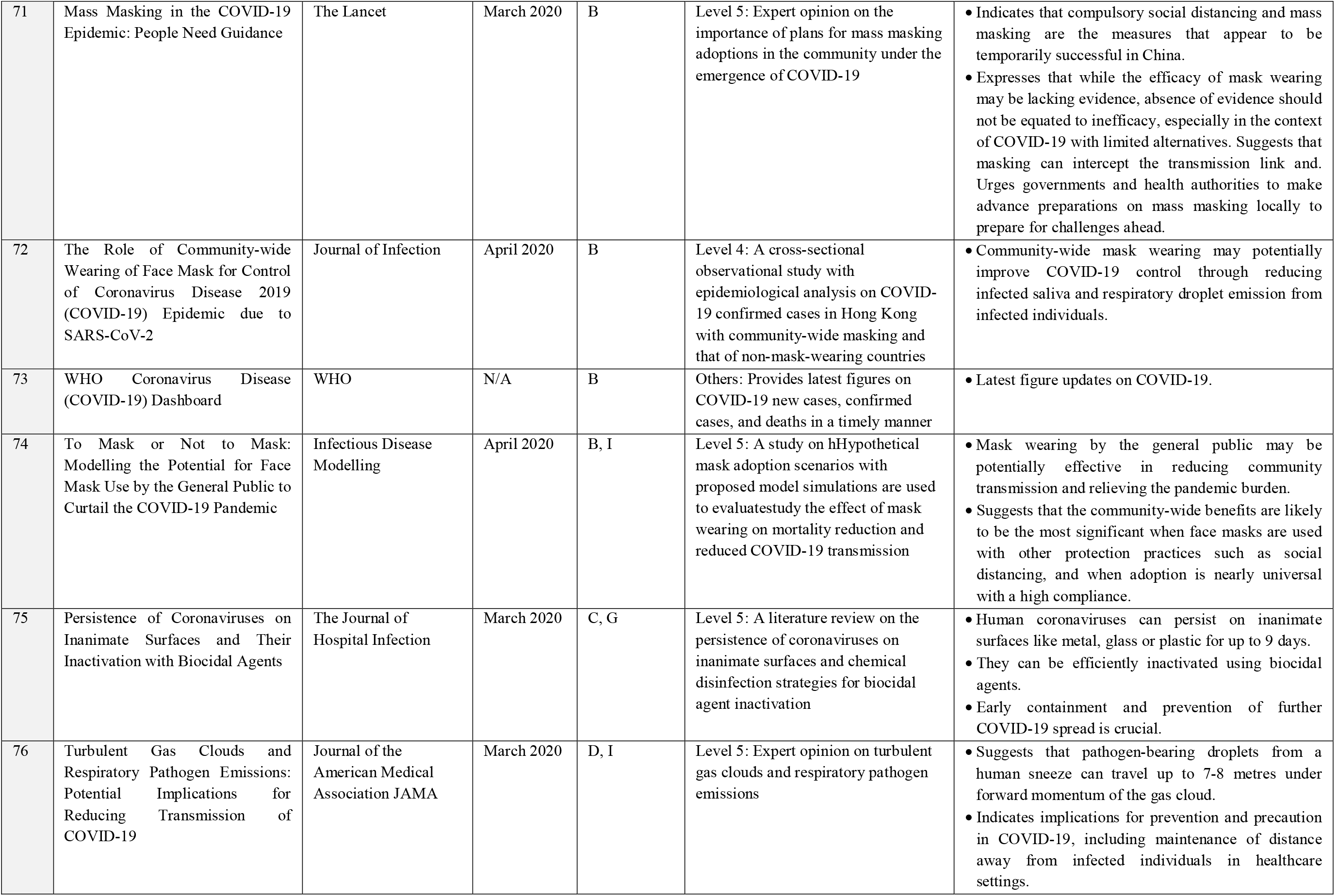
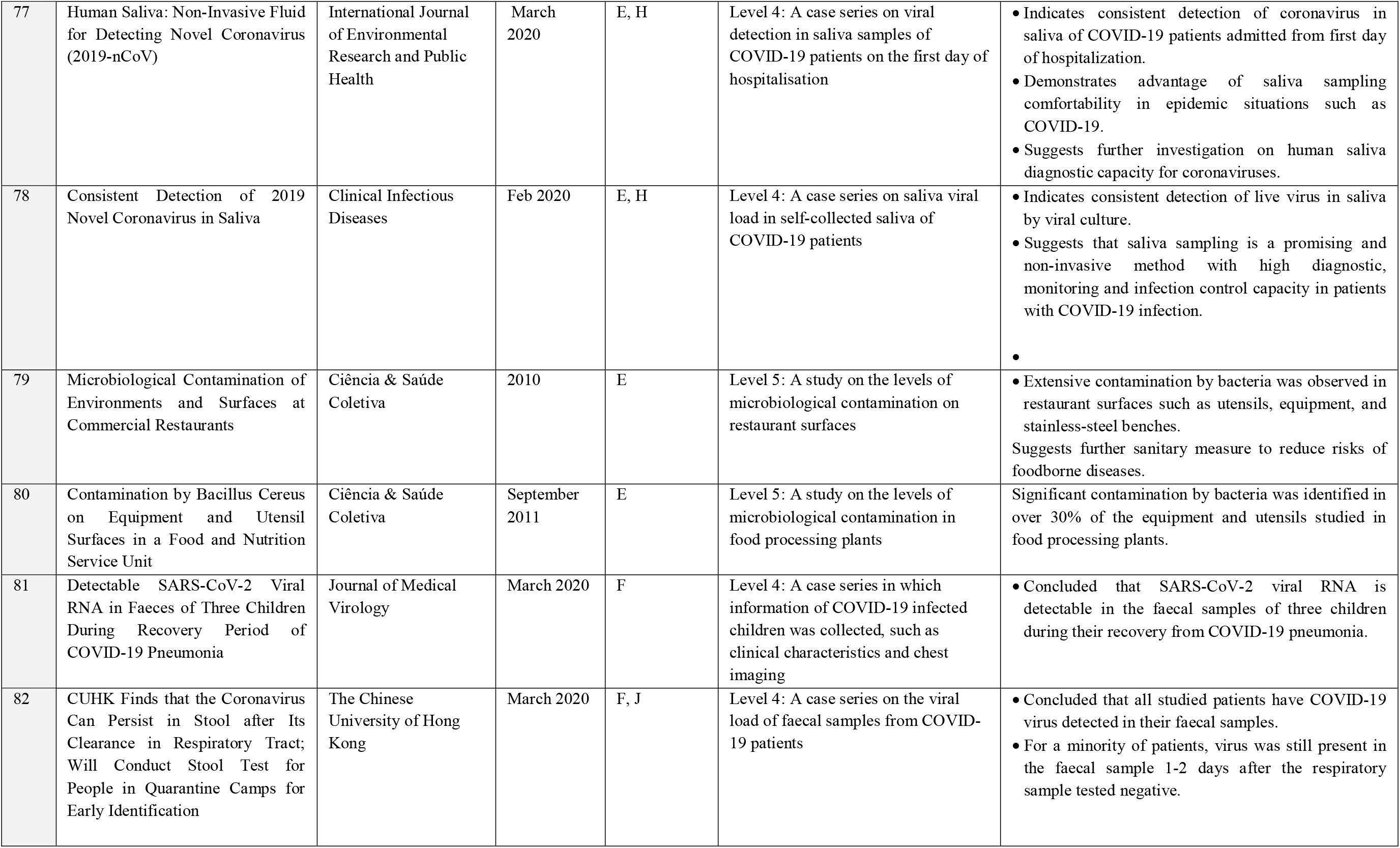
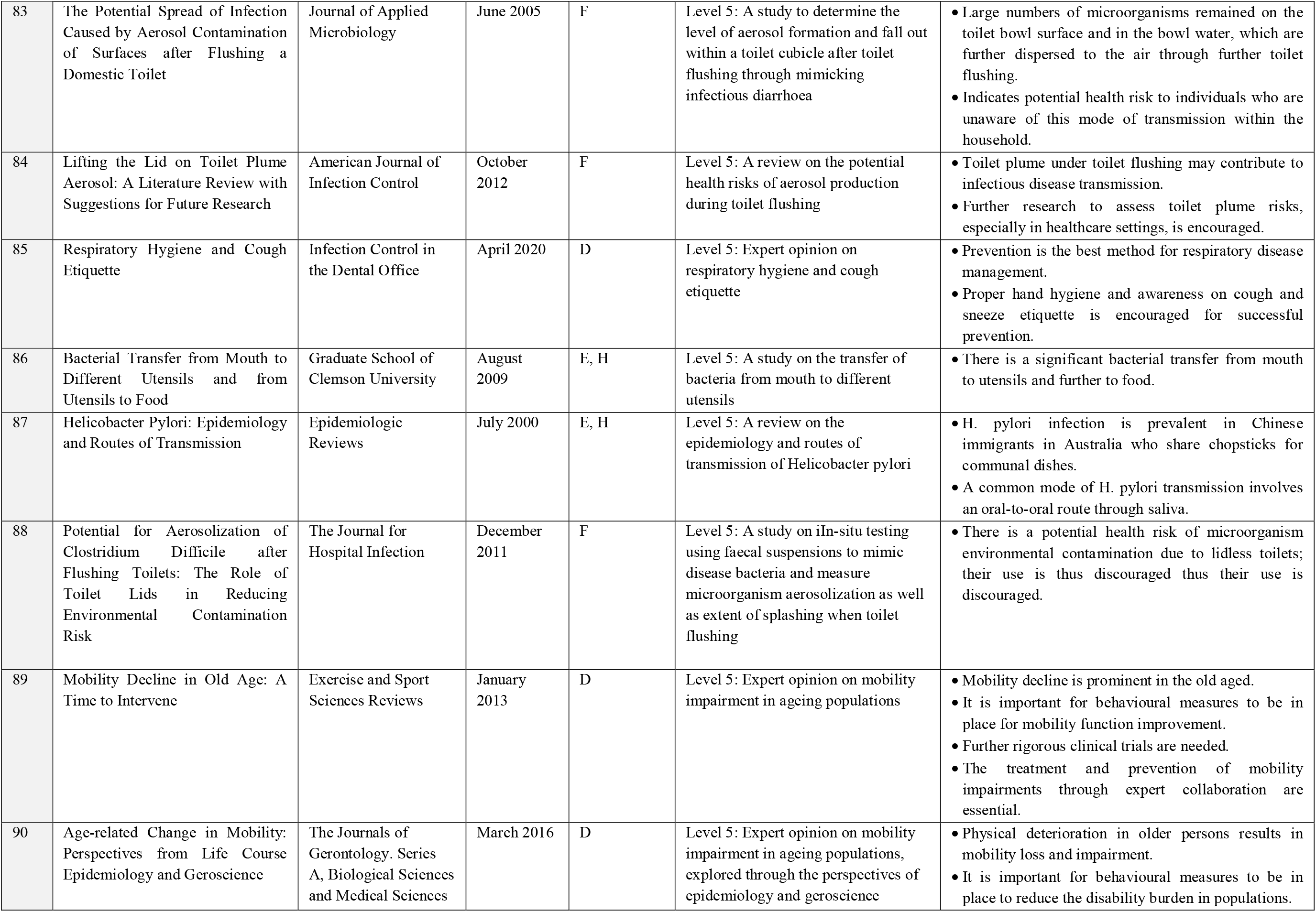
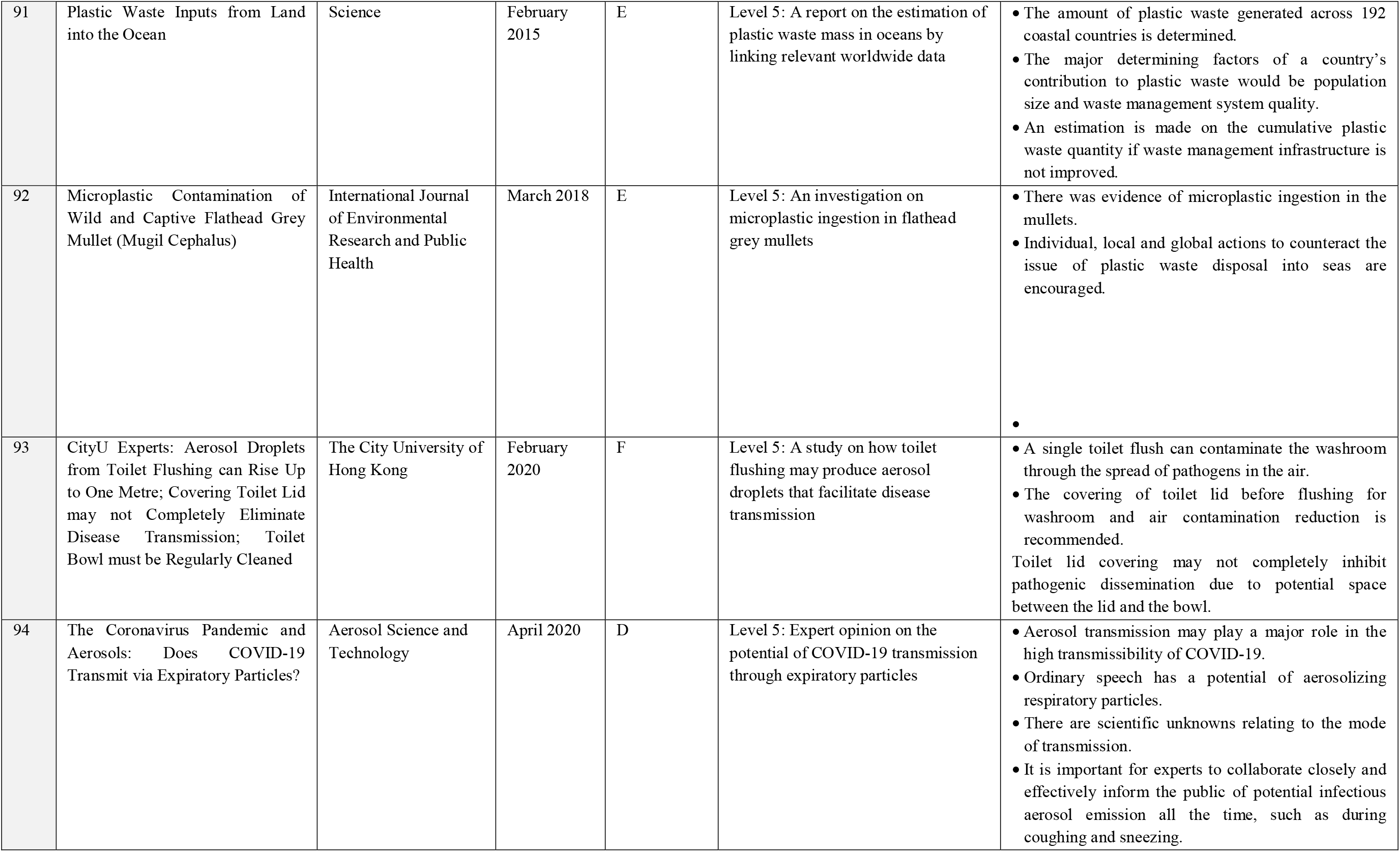
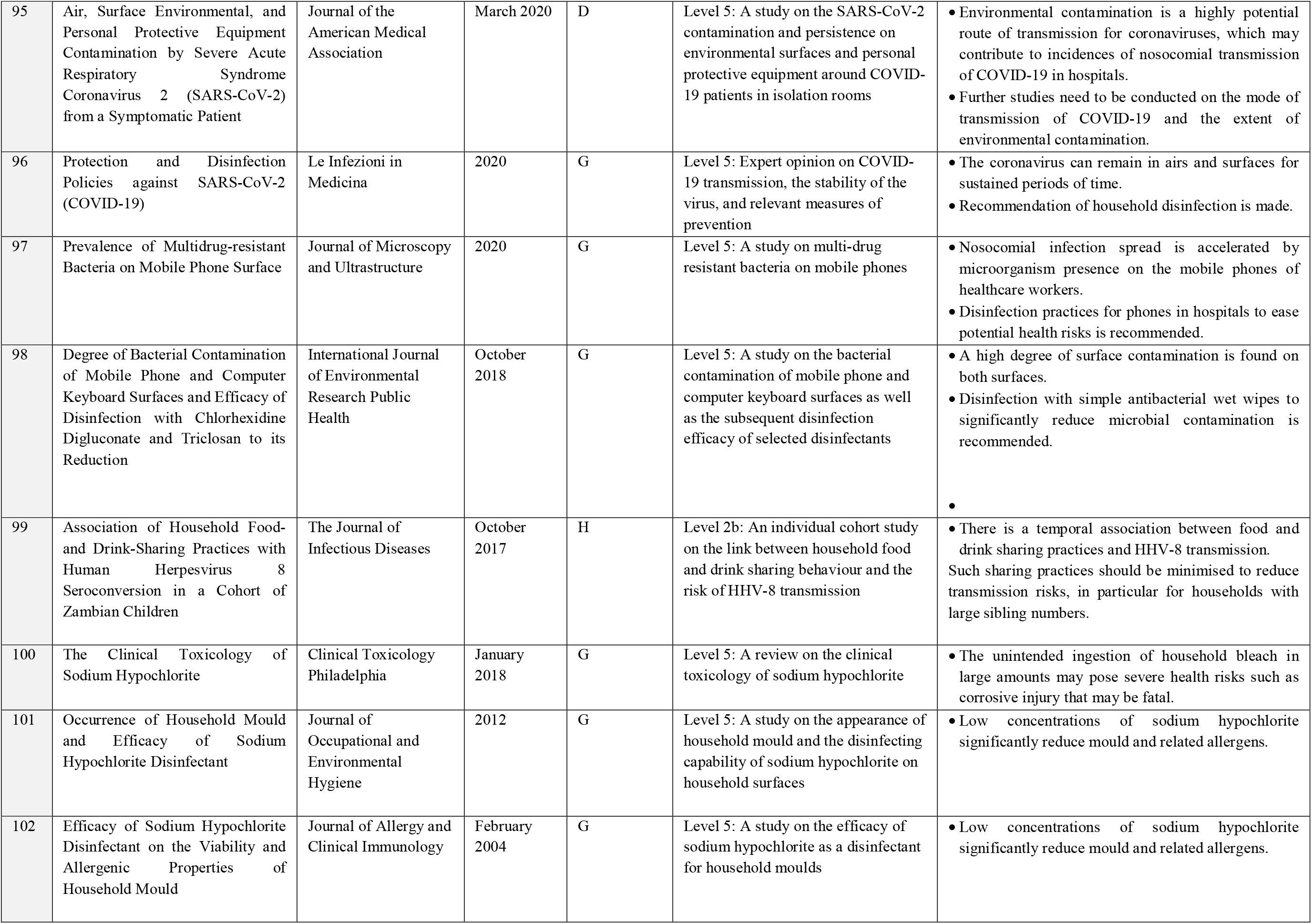
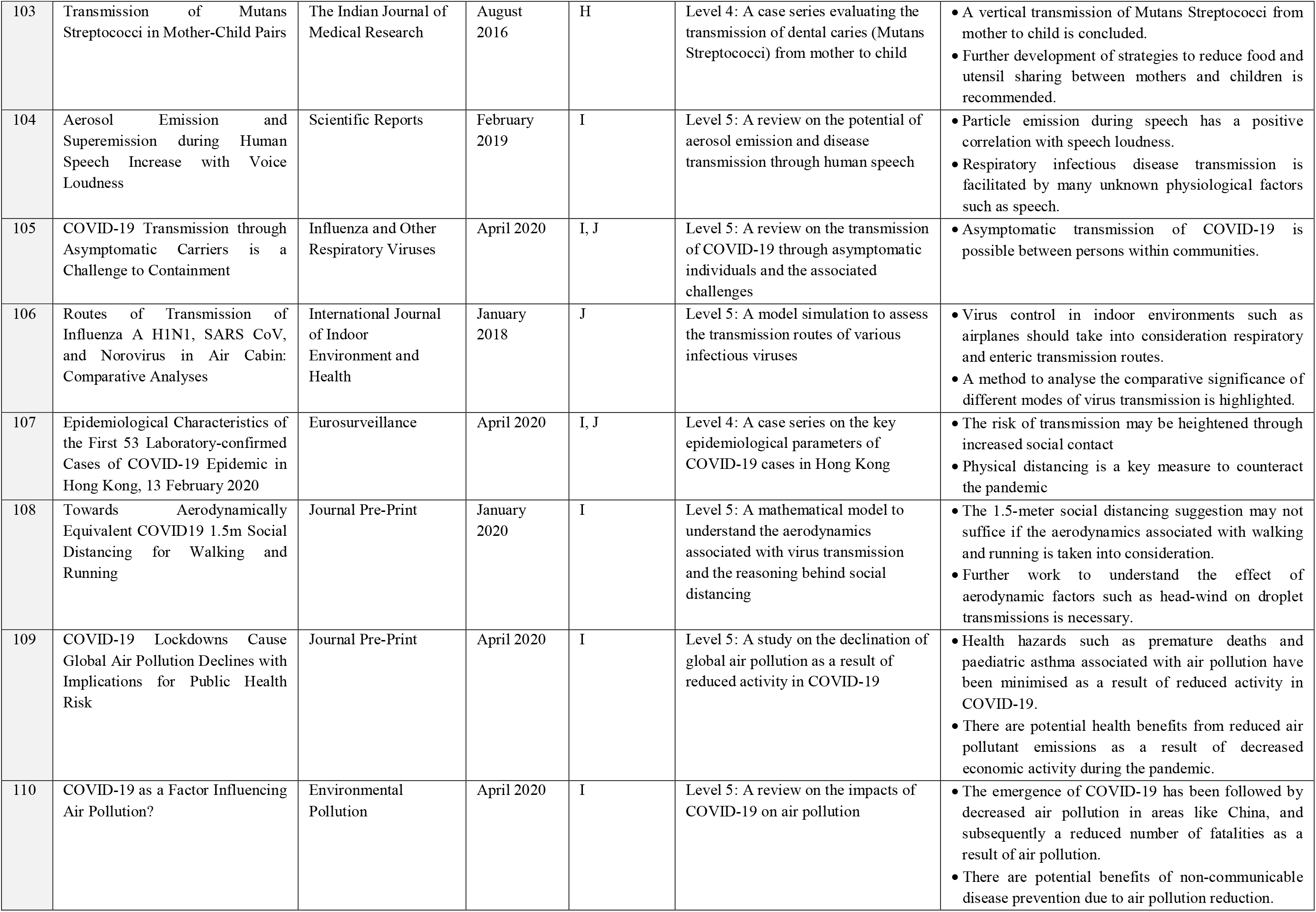
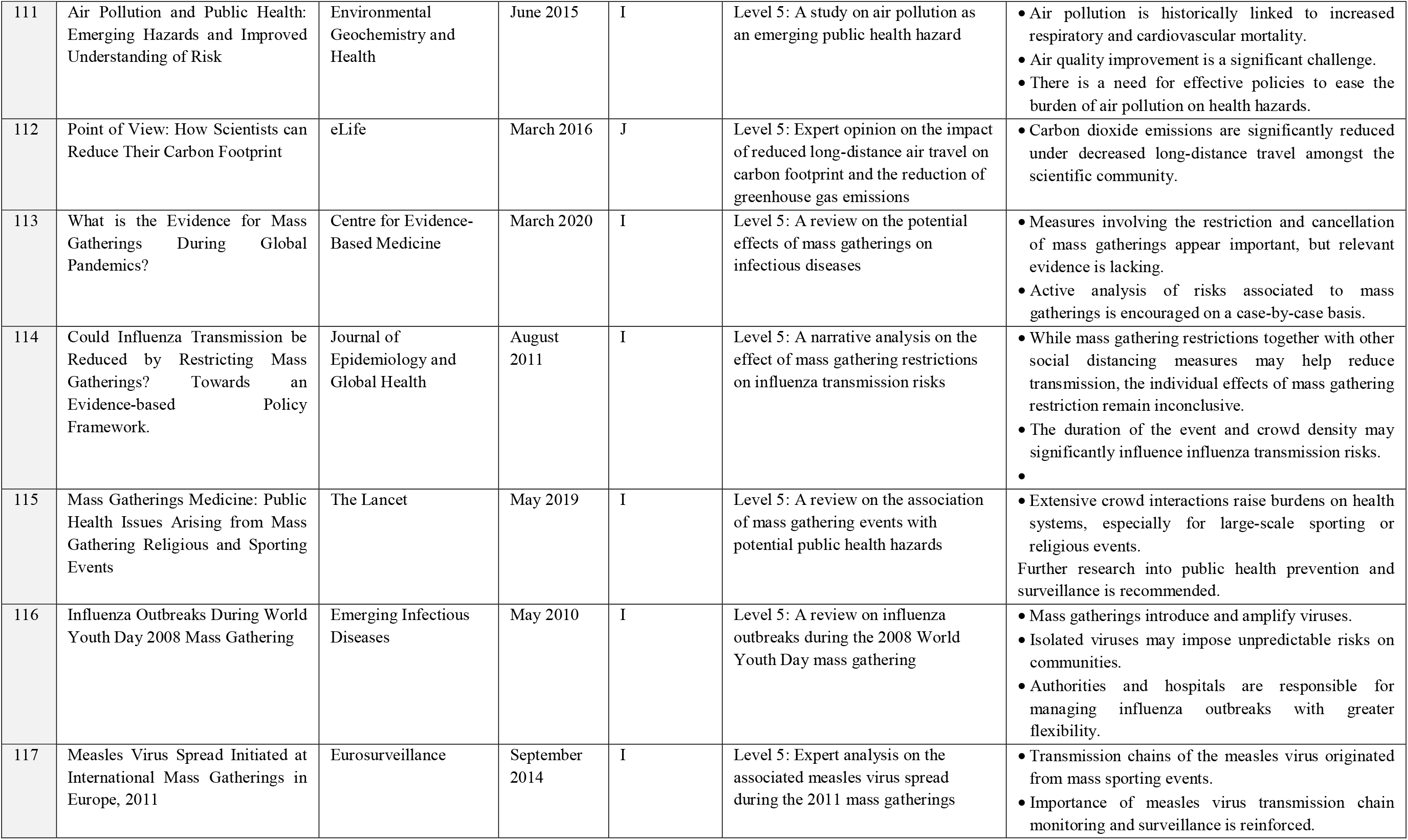

Appendix 1: Relevant Measure(s), Study Design, Relevant Key Finding(s) and/or Conclusion of Each Utilised Reference
| Ref. No. | Title | Journal or Publication | Date of Publication | Relevant Measure(s) (See Key 1) | OCEBM Level of Evidence based on Study Design (See Key 2) | Relevant Key Finding(s) and/or Conclusion |
| --- | --- | --- | --- | --- | --- | --- |
| 4 | Modes of Transmission of Virus Causing COVID-19: Implications for IPC Precaution Recommendations | WHO Scientific Brief | March 2020 | A, B, I | Level 5: Expert opinion on precaution recommendations, using research on the characteristics of COVID-19 | • With knowledge of droplet transmission (and particle size), droplet and contact precautions are recommended for COVID-19.<br>• Importance of PPE and other practices such as frequent hand hygiene is indicated. |
| 14 | COVID-19: A Call for Physical Scientists and Engineers | American Chemical Society NANO | April 2020 | A, D, H, I | Level 5: Expert opinion based on clinicians' experiences and knowledge; presentation of questions, hypotheses and research needs regarding COVID-19 | • Elucidates basic biology of viruses and their transmission and infection pathway.<br>• Importance of handwashing and hygiene is demonstrated via explanation of the need to deactivate released virions before they reach a host.<br>• Identifies the major complications and understandings associated with current measures such as PPE and surface sanitization and make recommendations accordingly. |
| 15 | Aerosol and Surface Stability of SARS-CoV-2 as Compared with SARS-CoV-1 | The New England Journal of Medicine | April 2020 | A, C, D, J | Level 5: An in vitro study of the surface stability of the SARS-Cov-2 strain compared to SARS-CoV-1 | • SARS-CoV-2 has similar surface stability compared to SARS-CoV-1 under experimental circumstances.<br>• Demonstrates stability on surfaces such as plastic and stainless steel with potential for aerosol and fomite transmission. |
| 16 | Community Transmission of Severe Acute Respiratory Syndrome Coronavirus 2, Shenzhen, China, 2020 | Emerging Infectious Diseases | June 2020 (Early Release) | B | Level 4: A case series on confirmed COVID-19 studied in order to understand the pattern of community transmission | • COVID-19 became endemic to Shenzhen. Community, intrafamily and nosocomial transmission-routes were found.<br>• Maintenance strategies are derived, such as minimising public activity, using personal protection measures and the importance of early screening, diagnosis and isolation. |
| 17 | A Familial Cluster of Pneumonia Associated with the 2019 Novel Coronavirus Indicating Person-to-Person Transmission: A Study of a Family Cluster | The Lancet | February 2020 | B | Level 4: A case series exploring epidemiological, clinical, laboratory, radiology and microbiological findings of a family cluster of (initially) unexplained pneumonia | • Indicating person-to-person transmission via nosocomial and intrafamily means.<br>• Noted that many findings were similar to those of SARS patients in 2003.<br>• One patient was initially asymptomatic – suggestion for early tracing, quarantine and control measures. |
| 18 | Tropism and Innate Host Responses of Influenza A/H5N6 Virus: An Analysis of Ex Vivo and In Vitro Cultures of the Human Respiratory Tract | European Respiratory Journal | March 2017 | B | Level 5: An in vitro study on tropism, replication competence and cytokine induction of virus isolates in cultures (Ex Vivo and In Vitro) derived from human respiratory tract | Human H5N6 virus adapted to human airways, indicating a of risk pattern for the virus upon entry into respiratory tract. |
| 19 | 2019-nCoV Transmission Through the Ocular Surface Must Not Be Ignored | The Lancet | February 2020 | B | Level 5: An ophthalmologist's expert perspective on additional risk through mucous membrane of eyes | Suggestion for consideration of studies into conjunctival scrapings to look for signs of ocular transmission.<br>Ophthalmologists must wear protective eyewear when examining suspect cases. |
| 20 | Presumed Asymptomatic Carrier Transmission of COVID-19 | Journal of the American Medical Association | February 2020 | A, B, D, I, J, H | Level 4: A case series on a familial cluster of 5 patients with COVID-19 | There is a potential mechanism of COVID-19 transmission via an asymptomatic carrier.<br>Further study on the relevant mechanism is suggested. |
| 21 | Emergencies Preparedness, Response: What Can I Do? | WHO | January 2010 | B | Level 5: A compilation of information on Pandemic Response (2009 H1N1) protective measures | Regarding masks specifically, it suggests that masks are only needed if you are sick.<br>Remarks on the importance of proper mask wearing practice if the measure is adopted. |
| 22 | SARS-CoV-2 Entry Factors are Highly Expressed in Nasal Epithelial Cells Together with Innate Immune Genes | Nature Medicine | April 2020 | B, C | Level 5: A study on SARS-CoV-2 tropism study via study of expression of viral entry-associated genes | Genes found to be co-expressed in nasal epithelial cells, indicating a role in the initial phase of viral infection, spread and clearance. |
| 23 | Use of Disposable Face Masks for Public Health Protection against SARS | Journal of Epidemiology and Community Health | April 2004 | A, B | Level 5: Expert opinion on the use of face masks and practice of personal hygiene as important measures to protect the general public from SARS | States that protection against SARS for healthcare workers is different from the general public, as the latter is not subject to continuous exposure to droplet transmission from an infected patient.<br>Expresses reduced risk of aerosol droplet transmission with masks.<br>Notes importance of proper usage and frequent changing of masks.<br>Also extends to mention importance of other personal hygiene practices such as handwashing due to survival of the virus on surfaces. |
| 24 | SARS Transmission, Risk Factors, and Prevention in Hong Kong | Emerging Infectious Diseases | April 2004 | A, B, G | Level 3b: A case-control study to compare SARS case-patients with undefined sources of infection with community controls | Concluded that risk factors for SARS infection include visiting mainland China, hospitals, and the Amoy Gardens (an estate with a SARS outbreak).<br>Indicates that frequent mask use in public venues, frequent handwashing, and household disinfection were prominent protective factors. |
| 25 | Respiratory Infections during SARS Outbreak, Hong Kong, 2003 | Emerging Infectious Diseases | November 2005 | A, B, D, G | Level 4: A cross-sectional study to compare the proportion of respiratory virus positive specimens in 2003 and those from 1998 to 2002 | • No direct causal relationship was established.<br>• However, the study suggests a positive association between reduced influenza/respiratory infection incidence and population-based hygienic measures including face mask wearing, hand washing after contact with potentially contaminated objects, using soap for handwashing, mouth covering when sneezing or coughing, and household disinfection. |
| 26 | Controlling the Novel A (H1N1) Influenza Virus: Don't Touch Your Face! | The Journal of Hospital Infection | November 2009 | A, C | Level 5: A letter to the editor on a study of surface swab specimens from patients with confirmed influenza A | • Indicates that virus strains of influenza A are found in surfaces such as bed rails, walls, and sofas.<br>• Further implies the importance of hand hygiene, droplet and contact precautions, and behavioural conditioning such as avoiding touching of the nose, eye or mouth touching to prevent and control influenza. |
| 27 | Stopping the Spread of COVID-19 | Journal of the American Medical Association | March 2020 | A, C, D, G, I | Level 5: A set of guidelines with potential measures to stop the spread of COVID-19 | • Different methods of infection prevention including hand hygiene, social distancing, household disinfection, and general personal hygiene are suggested. |
| 28 | Prevention of Coronavirus Disease 2019 (COVID-19) | Hong Kong Centre for Health Protection | May 2020 | A, B, C, D, F, G, I, J | Level 5: A set of guidelines with information related to COVID-19 such as prevention suggestions and clinical features of the coronavirus. | • Prevention advice such as mask wearing, avoidance of face touching, covering mouth and nose, putting the toilet lid down when flushing, and general travel advice is suggested. |
| 29 | Fact Sheet for SARS Patients and Their Close Contact | Centres for Disease Control and Prevention | 2003 | A, B, C, D, E, F, H, I, J | Level 5: A set of guidelines with information related to SARS such as symptoms, mode of transmission, and prevention measures | • Personal protection measures, such as the avoidance of silverware sharing, handwashing and covering mouth and nose when coughing or sneezing, are recommended. |
| 30 | WHO Guidelines on Hand Hygiene in Health Care | WHO | 2009 | A | Level 5: An extensive evidence-based guideline on the practice and science behind handwashing | • Extensive findings on best handwashing practice and efficacy of soap-based washing and alcohol against enveloped viruses.<br>• |
| 31 | Simplifying the World Health Organization Protocol: 3 Steps Versus 6 Steps for Performance of Hand Hygiene in a Cluster-randomized Trial | Clinical Infectious Diseases | August 2019 | A | Level 1b: A cluster-randomized trial assigning 3-step vs. 6-step handwashing protocol | • Findings suggest that both significantly reduced the bacterial colony (with no significant difference between the two) but that the 3-step guidelines had higher compliance.<br>Quantity of steps are not of great concern as long as areas are covered. |
| 32 | The Common Missed Handwashing Instances and Areas after 15 Years of Hand-Hygiene Education | Journal of Environmental and Public Health | August 2019 | A | Level 4: A cross-sectional study looking at a cohort in Hong Kong and their handwashing and hand hygiene practices | Indicates several areas of the hands which are commonly missed, as well as occasions during which handwashing should be performed.<br>Relationship between age or education and hand hygiene practice is indicated. |
| 33 | Hygiene and Health: Systematic Review of Handwashing Practices Worldwide and Update of Health Effects | Tropical Medicine and International Health | May 2014 | A | Level 1a: A systematic review of RCT's and quasi-randomised trials (+others). Studies observed rates of handwashing with soap in various populations and scenarios | Significant global problem regarding poor practice of handwashing after contact with excrete is found. |
| 34 | Assessment of Hand Hygiene Techniques Using the World Health Organization's Six Steps | Journal of Infection and Public Health | Dec 2015 | A | Level 2b: An individual cohort study observing hand hygiene techniques over a period of 5 months | Certain areas of the hand achieved lower areas of compliance during handwashing. |
| 35 | Bacteriological Aspects of Hand Washing: A Key for Health Promotion and Infections Control | International Journal of Preventative Medicine | March 2017 | A | Level 3a: A systematic review of case control studies | Handwashing can reduce infectious agent's transmission in the community and healthcare settings. |
| 36 | Coronavirus Disease (COVID-19) Advice for the Public | WHO | April 2020 | A, B, C, D, I | Level 5: Expert opinion on personal protection from COVID-19 such as safe use of alcohol-based hand sanitizers | Informs the public of the importance of actions such as regular handwashing with soap and water, cleaning hands with alcohol-based rub, social distancing, avoiding crowds, avoid eye, nose, mouth touching, covering mouth and nose, staying home, and health-seeking behaviour under the pandemic.<br>Precautions on alcohol-based hand sanitizer use is also mentioned. |
| 37 | How to Protect Yourself & Others | Centres for Disease Control and Prevention | April 2020 | A, B, D, G, I | Level 5: Expert opinion on how COVID-19 spreads and personal protection measures for COVID-19 | Informs the public of person-to-person spread of the virus, the lack of vaccine to prevent COVID-19, and the importance of actions such as regular handwashing with soap and water, avoiding close contact, covering mouth and nose with a cloth face cover, covering coughs and sneezes, as well as cleaning and disinfecting frequently touched surfaces and households under the pandemic. |
| 38 | Hand Hygiene and the Novel Coronavirus Pandemic: The Role of Healthcare Workers | The Journal of Hospital Infection | March 2020 | A | Level 5: Expert opinion on the importance of practicing respiratory and hand hygiene, as well as using personal protective equipment in healthcare settings | Details the role of healthcare workers, nurses and midwives in providing primary point of care in communities and for pregnant women respectively, especially during infectious disease outbreaks.<br>Mentions details and precautions when using alcohol-based hand rubs for hand hygiene. |
| 39 | A Schlieren Optical Study of the Human Cough With and Without Wearing Masks for Aerosol Infection Control | Journal of the Royal Society, Interface | December 2009 | B, D | Level 5: A study comparing the fluid dynamics of coughing with or without standard surgical or N95 mask wearing using video records. | • Human coughing projects a rapid turbulent jet into the surrounding air<br>• Wearing a surgical or N95 mask interrupts the natural mechanism of airborne infection transmission through blocking turbulent jet formation (N95 mask) or redirecting the exhalant (surgical mask). |
| 40 | Respiratory Virus Shedding in Exhaled Breath and Efficacy of Face Masks | Nature Medicine | April 2020 | B | Level 1b: A randomized controlled trial comparing exhaled breath samples (for respiratory virus shedding) in mask wearing vs. non mask wearing individuals | • Surgical face masks can prevent transmission of human coronaviruses and influenza viruses from symptomatic individuals.<br>• Surgical face masks reduce detection of coronavirus RNA in aerosols, with a trend towards reduced detection of coronavirus RNA in respiratory droplets. |
| 41 | Early Transmission Dynamics in Wuhan, China, of Novel Coronavirus-infected Pneumonia | The New England Journal of Medicine | January 2020 | B | Level 4: A case series looking at characteristics and illness timelines of laboratory confirmed cases of COVID-19 | • Human-to-human transmission has occurred and that measures must be implemented towards populations at risk. |
| 42 | Contamination by Respiratory Viruses on Outer Surface of Medical Masks Used by Hospital Healthcare Workers | BMC Infectious Diseases | June 2019 | B | Level 1b: An individual randomised controlled trial with two pilot studies (cohort). Participants told to wear medical masks and then masks were checked for respiratory viruses on the surface | • Virus presence on the face mask was higher when worn for a longer period of time (in the 6> subgroup).<br>• The study concluded that because of this risk, the pathogens on the outer surface may cause self-contamination, with greater risk when worn for >6 hours.<br>• Indications that there should be a maximum time on mask usage. |
| 43 | Stability of SARS-CoV-2 in different environmental conditions | The Lancet | May 2020 | B, G | Level 5: An experimental study on the stability of COVID-19 in different induced environmental conditions such as under heat stress and on different surfaces | • Infectious virus was not detected after 5 minutes incubation at room temperature, wide range of pH stability, was found to be stable on surfaces such as the outer layer of surgical masks.<br>• Virus was susceptible to disinfection methods. |
| 44 | What You Need to Know About Infectious Disease | US Institute of Medicine | 2010 | C, D | Level 5: A book that contains expert opinion on infectious diseases and the nature of their transmission | • The mouth, the eyes and the nose are the body's main entry points for transmittable conditions such as influenza.<br>• Coughing and sneezing facilitates the spread of droplet-transmittable diseases. |
| 45 | Face Touching: A Frequent Habit that Has Implications for Hand Hygiene | American Journal of Infection Control | February 2015 | C | Level 2b: A behavioural observation study of 26 participants exploring the habit of face touching | • Even amongst medical students, there was frequent face touching behaviour.<br>• This indicates towards the importance of hand hygiene too apart from the risk of self-inoculation from face touching which needs to be elucidated. |
| 46 | Self-touch: Contact Durations and Point of Touch of Spontaneous Facial Self-touches Differ Depending on Cognitive and Emotional Load | PLOS ONE Medicine (Baltimore) | March 2019 | C | Level 2b: A cohort study exploring the behaviour of face touching and its link to cognitive and emotional load | • Results showed the both the point of touch and contact durations were under influence from emotional and cognitive triggers. |
| 47 | Protective Effect of Hand-washing and Good Hygienic Habits against Seasonal Influenza: A Case-control Study | Medicine (Baltimore) | March 2016 | A | Level 3b: A single case-control study testing the link between influenza transmission and self-reported handwashing / unhealthy hygiene habits | • Frequent handwashing and better hygiene habits were associated with a reduction in the risk of influenza infection. |
| 48 | Hand Hygiene and Risk of Influenza Virus Infections in the Community: A Systematic Review and Meta-analysis | Epidemiology & Infection | May 2014 | A | Level 1a: A systematic review of 10 randomized controlled trials aiming to evaluate the efficacy of hand hygiene measures against the reduction of influenza transmission | • Findings suggested that while hand washing may be effective (modest efficacy) against one mode of transmission i.e. contact, further measures may also be important to control influenza transmission, for example face masks. |
| 49 | Effect of Washing Hands with Soap on Diarrhoea risk in the community: A Systematic Review | Cochrane Database of Systematic Reviews | September 2015 | A | Level 1a: A systematic review of 22 randomised controlled trials to compare diarrhoea occurrence in children and adults with or without handwashing measures | • Handwashing measures result in diarrhoea episode reductions in child day-care centres in high-income countries as well as communities in low- and middle-income countries.<br>• It is a challenge to encourage the habitual maintenance of handwashing habits in people in the long-term. |
| 50 | Hand Washing Promotion for Preventing Diarrhoea | Cochrane Systematic Review | September 2015 | A | Level 1a: A systematic review of randomized controlled trials and cluster RCT's to compare the effects of measures associated with handwashing on the occurrence of diarrhoea episodes in children | • Hand washing most likely reduces diarrhoea episodes in certain communities, as per the study's findings.<br>• There may be lack of understanding on how to help people maintain habits related to handwashing in the long term. |
| 51 | Reducing the Risk of Infection: Hand Washing Technique | Community Eye Health | March 2008 | A | Level 5: Expert guidance on components of a good handwashing route | Indicates that handwashing is critical to infection control and that there may be inadequate awareness on importance of handwashing techniques, which may be impeding effectiveness. |
| 52 | The Effectiveness of Hand Hygiene Procedures in Reducing the Risks of Infections in Home and Community Settings Including Handwashing and Alcohol-based Hand Sanitizers | American Journal of Infection Control | December 2007 | A | Level 5: A report reviewing the evidence on hand hygiene and its link to infectious disease transmissions | • Hand hygiene is a significant component of good hygiene in households and communities and has significant benefit towards the reduction of infection transmission, including respiratory tract infections.<br>• Further conclusion that hand hygiene's impact towards infectious disease reduction can be enhanced by improved persuasion of community handwashing (properly and at the right times) and that hand hygiene promotion should come hand in hand with other aspects of hygiene and associated education.<br>• |
| 53 | Effectiveness of Commercial Face Masks to Reduce Personal PM Exposure. | Science of the Total Environment | September 2018 | B | Level 5: A model-based study evaluating the efficacy of face mask respirators towards the reduction of airborne particle exposure and subsequent pollutant exposure | • Facemasks reduce exposure to urban pollution. The efficacy of available face masks can vary in achieving exposure reduction to urban pollution. |
| 54 | Exploring Motivations behind Pollution-mask Use in a Sample of Young Adults in Urban China | Globalization and Health | December 2018 | B | Level 4: A cross-sectional survey exploring the role of socio-cognitive factors in affecting the decision of wearing a pollution mask in the context of young educated people | • Mask wearing practice is influenced by various reasons including but not limited to level of education, social norms, self-efficacy, attitudes and past behaviour.<br>• The conclusion indicates the need towards changing the social perception towards mask-wearing practice. |
| 55 | WHO \| Air Pollution | WHO | N/A | B, I | Level 5: A collection of resources including global data on air pollution and subsequent protective measures | • Demonstrates that 9/10 people breathe air containing high levels of pollutants and concludes these as risk factors towards health. |
| 56 | Air Pollution: A Smoking Gun for Cancer | Chinese Journal of Cancer | April 2014 | B | Level 5: A review on various articles to discuss key questions surrounding the link of air pollution with cancer incidence, with a focus on China | • Air pollution was and is a risk for cancer; makes final recommendations such as the need for personal pollution monitoring devices as well as increase international collaborations upon this matter. |
| 57 | A Retrospective Approach to Assess Human Health Risks Associated with Growing Air Pollution in Urbanized Area of Thar Desert, Western Rajasthan, India | Journal of Environmental Health Science and Engineering. | January 2014 | B | Level 2b: A retrospective cohort study looking into the air pollution measures and associated statistics on disease burden | • Environmental burden of disease and association to air pollution is a main concern in the fast-developing areas of India.<br>• Households exposed to high vehicle-caused pollution presented with greater prevalence of respiratory diseases for example. |
| 58 | Saliva and Viral Infections | Periodontology 2000 | December 2015 | C, E | Level 5: A review on various publications associated with viral infections via the oral cavity and discussing assays | • Regarding saliva and its role in viral infections, it indicates that it plays a key role and that the mouth and eye are common sites for viral entry.<br>• Conclusion that the oral cavity is a significant area for infection as well as virus transmission. |
| 59 | Detection of Bacterial Pathogens in the Hands of Rural School Children Across Different Age Groups and Emphasizing the Importance of Hand Wash | Journal of Preventive Medicine and Hygiene | June 2019 | C | Level 4: A cross-sectional observational study on hand pathogens in 200 rural school children | • Concluded that in this rural-based cohort, the hands of the children were harbouring various, potentially fatal, pathogenic organisms and could thus be a major source of infection.<br>• Indication towards the importance of hand washing and the need to provide materials (which are not available to these groups) in order to reduce spread of infection which is otherwise reducible via hand hygiene. |
| 60 | Viricidal Activity of World Health Organization-recommended Formulations Against Enveloped Viruses, Including Zika, Ebola, and Emerging Coronaviruses | The Journal of Infectious Diseases | March 2017 | A | Level 5: An in vitro experiment testing the efficacy of two WHO recommended alcohol-based formulations against different enveloped viruses | • WHO recommended alcohol-based formulations worked against the different enveloped viruses and the viricidal effect was strong. |
| 61 | Effect of Handwashing on Child Health: A Randomised Controlled Trial | The Lancet | July 2005 | A | Level 1b: A randomised controlled trial randomly assigning of handwashing promotion to one group and no promotion to the other vs. randomised controls. Outcomes explored included diarrhoea and acute respiratory tract infections. | • Study found that households receiving plain soap with handwashing promotion had lower incidence of the studied infections and that there was not much difference between plain vs. antibacterial soap.<br>• Indicates the importance of such programs and distribution of soap.<br>• Concluding that handwashing was effective in preventing conditions like diarrhoea and respiratory disease. |
| 62 | Hand Cleaning with Ash for Reducing the Spread of Viral and Bacterial Infections: A Rapid Review | Cochrane | April 2020 | A | Level 5: A systematic review using different types of studies to assess the advantages and disadvantages of ash as an alternative to soap or other materials against viruses and bacteria | • Studies were unreliable and rarely adequate examined rate of infection. Therefore, ash could not be concluded as a suitable alternative. |
| 63 | Comparison of Four Methods of Hand Washing in Situations of Inadequate Water Supply | West African Journal of Medicine | January 2008 | A | Level 1b: A randomised controlled trial comparing different methods of hand washing developed for use in developing countries | • The 'Elbow way' of handwashing is the gold standard with no evidence of post-contamination.<br>• Bucket and bowl as well as the single-bowl method result in cross contamination. |
| 64 | Testing the Efficacy of Homemade Masks: Would They Protect in an Influenza Pandemic? | Disaster Medicine and Public Health Preparedness | August 2013 | A | Level 1b: A randomised controlled trial on the effectiveness of different household materials in making homemade masks as an alternative to commercial face masks | • While a homemade mask also results in a decrease in number of microorganisms expelled by volunteers, a homemade mask is significantly less effective than surgical masks and should only be a last resort for droplet transmission prevention. |
| 65 | Aerosol Filtration Efficiency of Common Fabrics Used in Respiratory Cloth Masks | American Chemical Society Nano | April 2020 | B | Level 5: An experimental approach to assess common fabrics (such as cotton) and their filtration efficiencies | • Found that in general, cloth masks could potentially offer notable protection against transmission of particles which have sizes within the aerosol range.<br>• Further findings on factors limiting effectiveness such as leakages due to fitting issues and influence of factors such as humidity, repeated use and washing.<br>• |
| 66 | Handwashing: Clean Hands Save Lives | Journal of Consumer Health on the Internet | February 2020 | A | Level 5: An expert collection of information on handwashing as well as the explanations behind it | Collects the key points on handwashing as well as the science behind the measure to ultimately make recommendations regarding when to wash and how to wash. |
| 67 | Effectiveness of Handwashing in Preventing SARS: A Review | Tropical Medicine and International Health | September 2006 | A | Level 3a: A systematic review of case-control studies to evaluate effectiveness of handwashing in protecting against SARS transmission | • Only three studies out of the ten reviewed were statistically significant.<br>• While there is no conclusive evidence on the effectiveness of handwashing, this measure remains suggestive to protect against SARS transmission in the community and healthcare settings. |
| 68 | Efficacy of Handwashing Duration and Drying Methods | International Association for Food Protection | July 2012 | A | Level 1b: A randomised controlled trial on the impact of soap or plain water, duration of practice, presence of debris and drying method on microorganism removal from hands through handwashing | • The use of soap, longer duration of handwashing, and towel drying significantly remove microorganisms compared to plain water, shorter duration and air drying respectively.<br>• Towel drying presented with a greater person-to-person variability. Presence of food debris made handwashing less effective. |
| 69 | Risk Factors for SARS among Persons without Known Contact with SARS Patients, Beijing, China | Emerging Infectious Diseases | February 2004 | B | Level 3b: An individual case-control study to compare unlinked probable SARS patients with other community-based controls | • Concluded that chronic medical conditions, visit to fever clinics, eating outside home, and frequent taxi taking were risk factors in case patients.<br>• Also indicated that mask wearing is strongly protective in reducing risk for SARS. |
| 70 | Effectiveness of Surgical and Cotton Masks in Blocking SARS-CoV-2: A Controlled Comparison in 4 Patients | Annals of Internal Medicine | April 2020 | B | Level 1b: A randomised controlled trial to compare the effectiveness of surgical and cotton masks in filtering SARS-CoV-2 | • Both surgical and cotton masks are potentially ineffective in the prevention of SARS-CoV-2 from patient coughs to the environment and external mask surface. |
| 71 | Mass Masking in the COVID-19 Epidemic: People Need Guidance | The Lancet | March 2020 | B | Level 5: Expert opinion on the importance of plans for mass masking adoptions in the community under the emergence of COVID-19 | • Indicates that compulsory social distancing and mass masking are the measures that appear to be temporarily successful in China.<br>• Expresses that while the efficacy of mask wearing may be lacking evidence, absence of evidence should not be equated to inefficacy, especially in the context of COVID-19 with limited alternatives. Suggests that masking can intercept the transmission link and. Urges governments and health authorities to make advance preparations on mass masking locally to prepare for challenges ahead. |
| 72 | The Role of Community-wide Wearing of Face Mask for Control of Coronavirus Disease 2019 (COVID-19) Epidemic due to SARS-CoV-2 | Journal of Infection | April 2020 | B | Level 4: A cross-sectional observational study with epidemiological analysis on COVID-19 confirmed cases in Hong Kong with community-wide masking and that of non-mask-wearing countries | • Community-wide mask wearing may potentially improve COVID-19 control through reducing infected saliva and respiratory droplet emission from infected individuals. |
| 73 | WHO Coronavirus Disease (COVID-19) Dashboard | WHO | N/A | B | Others: Provides latest figures on COVID-19 new cases, confirmed cases, and deaths in a timely manner | • Latest figure updates on COVID-19. |
| 74 | To Mask or Not to Mask: Modelling the Potential for Face Mask Use by the General Public to Curtail the COVID-19 Pandemic | Infectious Disease Modelling | April 2020 | B, I | Level 5: A study on hHypothetical mask adoption scenarios with proposed model simulations are used to evaluate the effect of mask wearing on mortality reduction and reduced COVID-19 transmission | • Mask wearing by the general public may be potentially effective in reducing community transmission and relieving the pandemic burden.<br>• Suggests that the community-wide benefits are likely to be the most significant when face masks are used with other protection practices such as social distancing, and when adoption is nearly universal with a high compliance. |
| 75 | Persistence of Coronaviruses on Inanimate Surfaces and Their Inactivation with Biocidal Agents | The Journal of Hospital Infection | March 2020 | C, G | Level 5: A literature review on the persistence of coronaviruses on inanimate surfaces and chemical disinfection strategies for biocidal agent inactivation | • Human coronaviruses can persist on inanimate surfaces like metal, glass or plastic for up to 9 days.<br>• They can be efficiently inactivated using biocidal agents.<br>• Early containment and prevention of further COVID-19 spread is crucial. |
| 76 | Turbulent Gas Clouds and Respiratory Pathogen Emissions: Potential Implications for Reducing Transmission of COVID-19 | Journal of the American Medical Association JAMA | March 2020 | D, I | Level 5: Expert opinion on turbulent gas clouds and respiratory pathogen emissions | • Suggests that pathogen-bearing droplets from a human sneeze can travel up to 7-8 metres under forward momentum of the gas cloud.<br>• Indicates implications for prevention and precaution in COVID-19, including maintenance of distance away from infected individuals in healthcare settings. |
| 77 | Human Saliva: Non-Invasive Fluid for Detecting Novel Coronavirus (2019-nCoV) | International Journal of Environmental Research and Public Health | March 2020 | E, H | Level 4: A case series on viral detection in saliva samples of COVID-19 patients on the first day of hospitalisation | • Indicates consistent detection of coronavirus in saliva of COVID-19 patients admitted from first day of hospitalization.<br>• Demonstrates advantage of saliva sampling comfortability in epidemic situations such as COVID-19.<br>• Suggests further investigation on human saliva diagnostic capacity for coronaviruses. |
| 78 | Consistent Detection of 2019 Novel Coronavirus in Saliva | Clinical Infectious Diseases | Feb 2020 | E, H | Level 4: A case series on saliva viral load in self-collected saliva of COVID-19 patients | • Indicates consistent detection of live virus in saliva by viral culture.<br>• Suggests that saliva sampling is a promising and non-invasive method with high diagnostic, monitoring and infection control capacity in patients with COVID-19 infection. |
| 79 | Microbiological Contamination of Environments and Surfaces at Commercial Restaurants | Ciência & Saúde Coletiva | 2010 | E | Level 5: A study on the levels of microbiological contamination on restaurant surfaces | • Extensive contamination by bacteria was observed in restaurant surfaces such as utensils, equipment, and stainless-steel benches.<br>Suggests further sanitary measure to reduce risks of foodborne diseases. |
| 80 | Contamination by Bacillus Cereus on Equipment and Utensil Surfaces in a Food and Nutrition Service Unit | Ciência & Saúde Coletiva | September 2011 | E | Level 5: A study on the levels of microbiological contamination in food processing plants | Significant contamination by bacteria was identified in over 30% of the equipment and utensils studied in food processing plants. |
| 81 | Detectable SARS-CoV-2 Viral RNA in Faeces of Three Children During Recovery Period of COVID-19 Pneumonia | Journal of Medical Virology | March 2020 | F | Level 4: A case series in which information of COVID-19 infected children was collected, such as clinical characteristics and chest imaging | • Concluded that SARS-CoV-2 viral RNA is detectable in the faecal samples of three children during their recovery from COVID-19 pneumonia. |
| 82 | CUHK Finds that the Coronavirus Can Persist in Stool after Its Clearance in Respiratory Tract; Will Conduct Stool Test for People in Quarantine Camps for Early Identification | The Chinese University of Hong Kong | March 2020 | F, J | Level 4: A case series on the viral load of faecal samples from COVID-19 patients | • Concluded that all studied patients have COVID-19 virus detected in their faecal samples.<br>• For a minority of patients, virus was still present in the faecal sample 1-2 days after the respiratory sample tested negative. |
| 83 | The Potential Spread of Infection Caused by Aerosol Contamination of Surfaces after Flushing a Domestic Toilet | Journal of Applied Microbiology | June 2005 | F | Level 5: A study to determine the level of aerosol formation and fall out within a toilet cubicle after toilet flushing through mimicking infectious diarrhoea | • Large numbers of microorganisms remained on the toilet bowl surface and in the bowl water, which are further dispersed to the air through further toilet flushing.<br>• Indicates potential health risk to individuals who are unaware of this mode of transmission within the household. |
| 84 | Lifting the Lid on Toilet Plume Aerosol: A Literature Review with Suggestions for Future Research | American Journal of Infection Control | October 2012 | F | Level 5: A review on the potential health risks of aerosol production during toilet flushing | • Toilet plume under toilet flushing may contribute to infectious disease transmission.<br>• Further research to assess toilet plume risks, especially in healthcare settings, is encouraged. |
| 85 | Respiratory Hygiene and Cough Etiquette | Infection Control in the Dental Office | April 2020 | D | Level 5: Expert opinion on respiratory hygiene and cough etiquette | • Prevention is the best method for respiratory disease management.<br>• Proper hand hygiene and awareness on cough and sneeze etiquette is encouraged for successful prevention. |
| 86 | Bacterial Transfer from Mouth to Different Utensils and from Utensils to Food | Graduate School of Clemson University | August 2009 | E, H | Level 5: A study on the transfer of bacteria from mouth to different utensils | • There is a significant bacterial transfer from mouth to utensils and further to food. |
| 87 | Helicobacter Pylori: Epidemiology and Routes of Transmission | Epidemiologic Reviews | July 2000 | E, H | Level 5: A review on the epidemiology and routes of transmission of Helicobacter pylori | • H. pylori infection is prevalent in Chinese immigrants in Australia who share chopsticks for communal dishes.<br>• A common mode of H. pylori transmission involves an oral-to-oral route through saliva. |
| 88 | Potential for Aerosolization of Clostridium Difficile after Flushing Toilets: The Role of Toilet Lids in Reducing Environmental Contamination Risk | The Journal for Hospital Infection | December 2011 | F | Level 5: A study on in-situ testing using faecal suspensions to mimic disease bacteria and measure microorganism aerosolization as well as extent of splashing when toilet flushing | • There is a potential health risk of microorganism environmental contamination due to lidless toilets; their use is thus discouraged thus their use is discouraged. |
| 89 | Mobility Decline in Old Age: A Time to Intervene | Exercise and Sport Sciences Reviews | January 2013 | D | Level 5: Expert opinion on mobility impairment in ageing populations | • Mobility decline is prominent in the old aged.<br>• It is important for behavioural measures to be in place for mobility function improvement.<br>• Further rigorous clinical trials are needed.<br>• The treatment and prevention of mobility impairments through expert collaboration are essential. |
| 90 | Age-related Change in Mobility: Perspectives from Life Course Epidemiology and Geroscience | The Journals of Gerontology. Series A, Biological Sciences and Medical Sciences | March 2016 | D | Level 5: Expert opinion on mobility impairment in ageing populations, explored through the perspectives of epidemiology and geroscience | • Physical deterioration in older persons results in mobility loss and impairment.<br>• It is important for behavioural measures to be in place to reduce the disability burden in populations. |
| 91 | Plastic Waste Inputs from Land into the Ocean | Science | February 2015 | E | Level 5: A report on the estimation of plastic waste mass in oceans by linking relevant worldwide data | • The amount of plastic waste generated across 192 coastal countries is determined.<br>• The major determining factors of a country's contribution to plastic waste would be population size and waste management system quality.<br>• An estimation is made on the cumulative plastic waste quantity if waste management infrastructure is not improved. |
| 92 | Microplastic Contamination of Wild and Captive Flathead Grey Mullet (Mugil Cephalus) | International Journal of Environmental Research and Public Health | March 2018 | E | Level 5: An investigation on microplastic ingestion in flathead grey mullets | • There was evidence of microplastic ingestion in the mullets.<br>• Individual, local and global actions to counteract the issue of plastic waste disposal into seas are encouraged.<br>• |
| 93 | CityU Experts: Aerosol Droplets from Toilet Flushing can Rise Up to One Metre; Covering Toilet Lid may not Completely Eliminate Disease Transmission; Toilet Bowl must be Regularly Cleaned | The City University of Hong Kong | February 2020 | F | Level 5: A study on how toilet flushing may produce aerosol droplets that facilitate disease transmission | • A single toilet flush can contaminate the washroom through the spread of pathogens in the air.<br>• The covering of toilet lid before flushing for washroom and air contamination reduction is recommended.<br>Toilet lid covering may not completely inhibit pathogenic dissemination due to potential space between the lid and the bowl. |
| 94 | The Coronavirus Pandemic and Aerosols: Does COVID-19 Transmit via Expiratory Particles? | Aerosol Science and Technology | April 2020 | D | Level 5: Expert opinion on the potential of COVID-19 transmission through expiratory particles | • Aerosol transmission may play a major role in the high transmissibility of COVID-19.<br>• Ordinary speech has a potential of aerosolizing respiratory particles.<br>• There are scientific unknowns relating to the mode of transmission.<br>• It is important for experts to collaborate closely and effectively inform the public of potential infectious aerosol emission all the time, such as during coughing and sneezing. |
| 95 | Air, Surface Environmental, and Personal Protective Equipment Contamination by Severe Acute Respiratory Syndrome Coronavirus 2 (SARS-CoV-2) from a Symptomatic Patient | Journal of the American Medical Association | March 2020 | D | Level 5: A study on the SARS-CoV-2 contamination and persistence on environmental surfaces and personal protective equipment around COVID-19 patients in isolation rooms | • Environmental contamination is a highly potential route of transmission for coronaviruses, which may contribute to incidences of nosocomial transmission of COVID-19 in hospitals.<br>• Further studies need to be conducted on the mode of transmission of COVID-19 and the extent of environmental contamination. |
| 96 | Protection and Disinfection Policies against SARS-CoV-2 (COVID-19) | Le Infezioni in Medicina | 2020 | G | Level 5: Expert opinion on COVID-19 transmission, the stability of the virus, and relevant measures of prevention | • The coronavirus can remain in airs and surfaces for sustained periods of time.<br>• Recommendation of household disinfection is made. |
| 97 | Prevalence of Multidrug-resistant Bacteria on Mobile Phone Surface | Journal of Microscopy and Ultrastructure | 2020 | G | Level 5: A study on multi-drug resistant bacteria on mobile phones | • Nosocomial infection spread is accelerated by microorganism presence on the mobile phones of healthcare workers.<br>• Disinfection practices for phones in hospitals to ease potential health risks is recommended. |
| 98 | Degree of Bacterial Contamination of Mobile Phone and Computer Keyboard Surfaces and Efficacy of Disinfection with Chlorhexidine Digluconate and Triclosan to its Reduction | International Journal of Environmental Research Public Health | October 2018 | G | Level 5: A study on the bacterial contamination of mobile phone and computer keyboard surfaces as well as the subsequent disinfection efficacy of selected disinfectants | • A high degree of surface contamination is found on both surfaces.<br>• Disinfection with simple antibacterial wet wipes to significantly reduce microbial contamination is recommended.<br>• |
| 99 | Association of Household Food- and Drink-Sharing Practices with Human Herpesvirus 8 Seroconversion in a Cohort of Zambian Children | The Journal of Infectious Diseases | October 2017 | H | Level 2b: An individual cohort study on the link between household food and drink sharing behaviour and the risk of HHV-8 transmission | • There is a temporal association between food and drink sharing practices and HHV-8 transmission. Such sharing practices should be minimised to reduce transmission risks, in particular for households with large sibling numbers. |
| 100 | The Clinical Toxicology of Sodium Hypochlorite | Clinical Toxicology Philadelphia | January 2018 | G | Level 5: A review on the clinical toxicology of sodium hypochlorite | • The unintended ingestion of household bleach in large amounts may pose severe health risks such as corrosive injury that may be fatal. |
| 101 | Occurrence of Household Mould and Efficacy of Sodium Hypochlorite Disinfectant | Journal of Occupational and Environmental Hygiene | 2012 | G | Level 5: A study on the appearance of household mould and the disinfecting capability of sodium hypochlorite on household surfaces | • Low concentrations of sodium hypochlorite significantly reduce mould and related allergens. |
| 102 | Efficacy of Sodium Hypochlorite Disinfectant on the Viability and Allergenic Properties of Household Mould | Journal of Allergy and Clinical Immunology | February 2004 | G | Level 5: A study on the efficacy of sodium hypochlorite as a disinfectant for household moulds | • Low concentrations of sodium hypochlorite significantly reduce mould and related allergens. |
| 103 | Transmission of Mutans Streptococci in Mother-Child Pairs | The Indian Journal of Medical Research | August 2016 | H | Level 4: A case series evaluating the transmission of dental caries (Mutans Streptococci) from mother to child | • A vertical transmission of Mutans Streptococci from mother to child is concluded.<br>• Further development of strategies to reduce food and utensil sharing between mothers and children is recommended. |
| 104 | Aerosol Emission and Superemission during Human Speech Increase with Voice Loudness | Scientific Reports | February 2019 | I | Level 5: A review on the potential of aerosol emission and disease transmission through human speech | • Particle emission during speech has a positive correlation with speech loudness.<br>• Respiratory infectious disease transmission is facilitated by many unknown physiological factors such as speech. |
| 105 | COVID-19 Transmission through Asymptomatic Carriers is a Challenge to Containment | Influenza and Other Respiratory Viruses | April 2020 | I, J | Level 5: A review on the transmission of COVID-19 through asymptomatic individuals and the associated challenges | • Asymptomatic transmission of COVID-19 is possible between persons within communities. |
| 106 | Routes of Transmission of Influenza A H1N1, SARS CoV, and Norovirus in Air Cabin: Comparative Analyses | International Journal of Indoor Environment and Health | January 2018 | J | Level 5: A model simulation to assess the transmission routes of various infectious viruses | • Virus control in indoor environments such as airplanes should take into consideration respiratory and enteric transmission routes.<br>• A method to analyse the comparative significance of different modes of virus transmission is highlighted. |
| 107 | Epidemiological Characteristics of the First 53 Laboratory-confirmed Cases of COVID-19 Epidemic in Hong Kong, 13 February 2020 | Eurosurveillance | April 2020 | I, J | Level 4: A case series on the key epidemiological parameters of COVID-19 cases in Hong Kong | • The risk of transmission may be heightened through increased social contact<br>• Physical distancing is a key measure to counteract the pandemic |
| 108 | Towards Aerodynamically Equivalent COVID19 1.5m Social Distancing for Walking and Running | Journal Pre-Print | January 2020 | I | Level 5: A mathematical model to understand the aerodynamics associated with virus transmission and the reasoning behind social distancing | • The 1.5-meter social distancing suggestion may not suffice if the aerodynamics associated with walking and running is taken into consideration.<br>• Further work to understand the effect of aerodynamic factors such as head-wind on droplet transmissions is necessary. |
| 109 | COVID-19 Lockdowns Cause Global Air Pollution Declines with Implications for Public Health Risk | Journal Pre-Print | April 2020 | I | Level 5: A study on the declination of global air pollution as a result of reduced activity in COVID-19 | • Health hazards such as premature deaths and paediatric asthma associated with air pollution have been minimised as a result of reduced activity in COVID-19.<br>• There are potential health benefits from reduced air pollutant emissions as a result of decreased economic activity during the pandemic. |
| 110 | COVID-19 as a Factor Influencing Air Pollution? | Environmental Pollution | April 2020 | I | Level 5: A review on the impacts of COVID-19 on air pollution | • The emergence of COVID-19 has been followed by decreased air pollution in areas like China, and subsequently a reduced number of fatalities as a result of air pollution.<br>• There are potential benefits of non-communicable disease prevention due to air pollution reduction. |
| 111 | Air Pollution and Public Health: Emerging Hazards and Improved Understanding of Risk | Environmental Geochemistry and Health | June 2015 | I | Level 5: A study on air pollution as an emerging public health hazard | • Air pollution is historically linked to increased respiratory and cardiovascular mortality.<br>• Air quality improvement is a significant challenge.<br>• There is a need for effective policies to ease the burden of air pollution on health hazards. |
| 112 | Point of View: How Scientists can Reduce Their Carbon Footprint | eLife | March 2016 | J | Level 5: Expert opinion on the impact of reduced long-distance air travel on carbon footprint and the reduction of greenhouse gas emissions | • Carbon dioxide emissions are significantly reduced under decreased long-distance travel amongst the scientific community. |
| 113 | What is the Evidence for Mass Gatherings During Global Pandemics? | Centre for Evidence-Based Medicine | March 2020 | I | Level 5: A review on the potential effects of mass gatherings on infectious diseases | • Measures involving the restriction and cancellation of mass gatherings appear important, but relevant evidence is lacking.<br>• Active analysis of risks associated to mass gatherings is encouraged on a case-by-case basis. |
| 114 | Could Influenza Transmission be Reduced by Restricting Mass Gatherings? Towards an Evidence-based Policy Framework. | Journal of Epidemiology and Global Health | August 2011 | I | Level 5: A narrative analysis on the effect of mass gathering restrictions on influenza transmission risks | • While mass gathering restrictions together with other social distancing measures may help reduce transmission, the individual effects of mass gathering restriction remain inconclusive.<br>• The duration of the event and crowd density may significantly influence influenza transmission risks.<br>• |
| 115 | Mass Gatherings Medicine: Public Health Issues Arising from Mass Gathering Religious and Sporting Events | The Lancet | May 2019 | I | Level 5: A review on the association of mass gathering events with potential public health hazards | • Extensive crowd interactions raise burdens on health systems, especially for large-scale sporting or religious events.<br>Further research into public health prevention and surveillance is recommended. |
| 116 | Influenza Outbreaks During World Youth Day 2008 Mass Gathering | Emerging Infectious Diseases | May 2010 | I | Level 5: A review on influenza outbreaks during the 2008 World Youth Day mass gathering | • Mass gatherings introduce and amplify viruses.<br>• Isolated viruses may impose unpredictable risks on communities.<br>• Authorities and hospitals are responsible for managing influenza outbreaks with greater flexibility. |
| 117 | Measles Virus Spread Initiated at International Mass Gatherings in Europe, 2011 | Eurosurveillance | September 2014 | I | Level 5: Expert analysis on the associated measles virus spread during the 2011 mass gatherings | • Transmission chains of the measles virus originated from mass sporting events.<br>• Importance of measles virus transmission chain monitoring and surveillance is reinforced. |

### KEY 1 – MEASURES

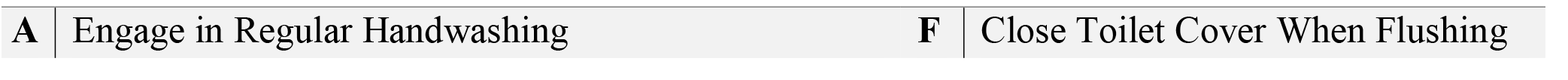
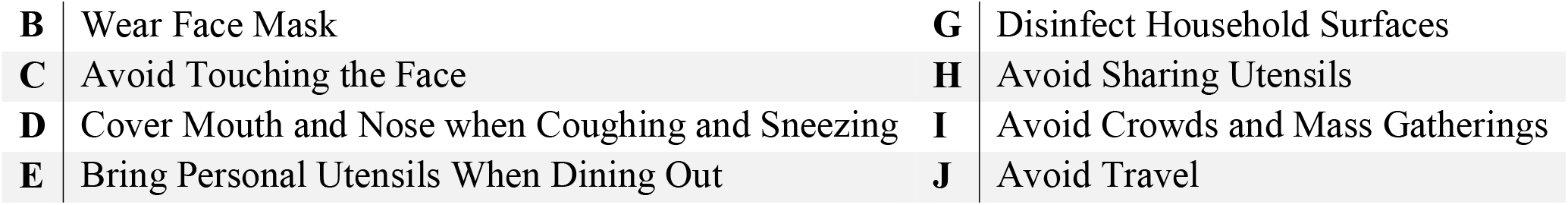

### KEY 2 – OCEBM LEVEL OF EVIDENCE

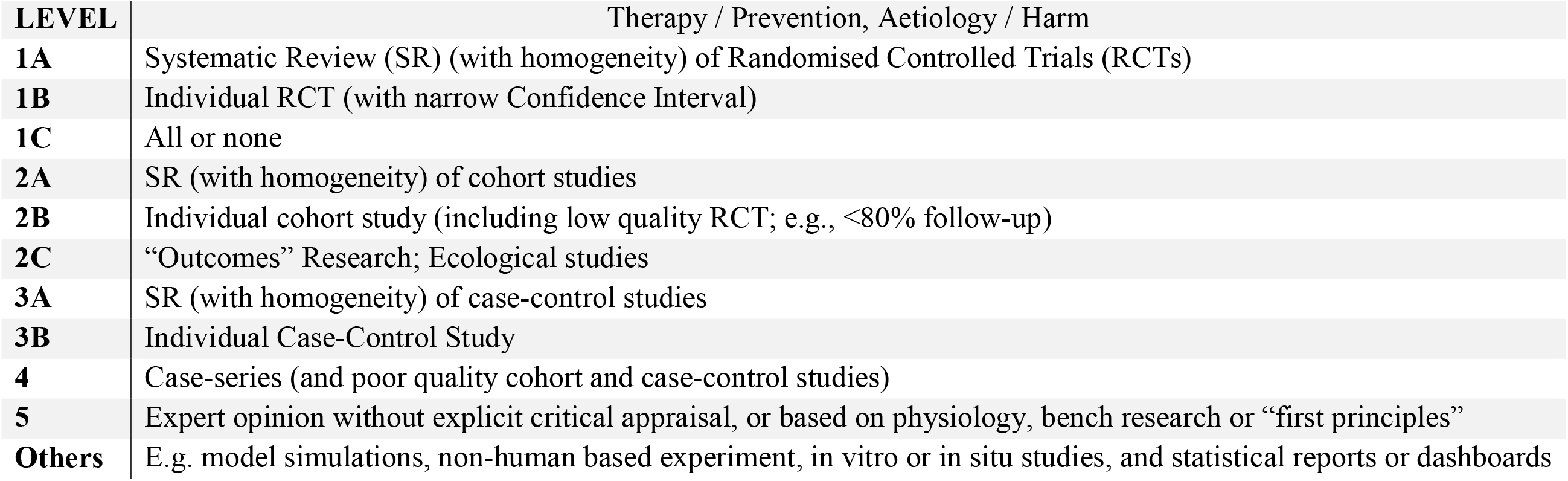

| LEVEL | Therapy / Prevention, Aetiology / Harm |
| --- | --- |
| **1A** | Systematic Review (SR) (with homogeneity) of Randomised Controlled Trials (RCTs) |
| **1B** | Individual RCT (with narrow Confidence Interval) |
| **1C** | All or none |
| **2A** | SR (with homogeneity) of cohort studies |
| **2B** | Individual cohort study (including low quality RCT; e.g., <80% follow-up) |
| **2C** | “Outcomes” Research; Ecological studies |
| **3A** | SR (with homogeneity) of case-control studies |
| **3B** | Individual Case-Control Study |
| **4** | Case-series (and poor quality cohort and case-control studies) |
| **5** | Expert opinion without explicit critical appraisal, or based on physiology, bench research or “first principles” |
| **Others** | E.g. model simulations, non-human based experiment, in vitro or in situ studies, and statistical reports or dashboards |

